# District-level HIV and TB health system vulnerability to climate and weather hazards in South Africa: a composite index approach

**DOI:** 10.64898/2026.08.21.26360993

**Authors:** Erofili Grapsa, Marlies Craig, Nondumiso Mthiyane, Sammy Khagayi, Saeideh Babashahi, Collins Iwuji, the ASTRA study group

**Affiliations:** Africa Health Research Institute, KwaZulu-Natal, South Africa; Department of Global Health and Infection, Brighton and Sussex Medical School, University of Brighton and University of Sussex, Brighton, UK; Dept of International Public Health, Liverpool School of Tropical Medicine, Liverpool, UK

## Abstract

**Background:** Climate change and extreme weather events (EWEs) threaten health systems, disrupt continuity of HIV and TB services, amplify communicable disease burdens, and exacerbate health inequities in South Africa. Yet few empirical studies have quantified district-level vulnerability, where HIV and TB service delivery and climate adaptation are operationalised.

**Methods:** We developed a district-level composite HIV/TB Vulnerability Index, integrating indicators of HIV & tuberculosis burden (sensitivity), health system capacity, and socio-economic vulnerability. We also developed a Hazard Index which when combined with the HIV/TB vulnerability Index, identifies districts where underlying vulnerability coincides with higher likelihood of EWEs. Indicators were drawn from national surveys, routine health information systems, and international hazard datasets, normalised using a min-max scaling, and aggregated with equal weighting. Sensitivity analysis were conducted to assess the robustness of the composite indices.

**Findings:** The most vulnerable districts were located in the Northern Cape, Eastern Cape and KwaZulu Natal provinces where high HIV/TB burden and socio-economic sensitivity coincided with limited health system adaptive capacity. In contrast, the least vulnerable districts, were concentrated in Gauteng and Western Cape, reflecting stronger health system capacity and more favourable socio-economic conditions. Hazard exposure exhibited a clear spatial division with western districts experiencing greater heat stress and eastern districts facing higher flood and heavy-rainfall hazards. When hazard exposure was combined with the HIV/TB vulnerability Index, districts with both high vulnerability and hazard scores clustered predominantly along the east coast (Ugu, uMkhanyakude, and Harry Gwala in KwaZulu-Natal, and O.R. Tambo and Alfred Nzo in the Eastern Cape).

**Interpretation:** South Africa’s district-level vulnerability to climate and weather hazards is driven by the convergence of high HIV/TB burden, constrained health system capacity, and socio-economic disadvantage. Where this vulnerability intersects with increased hazard risk, it creates a compound susceptibility that needs attention. Our findings provide evidence for geographically targeted adaptation, prioritising continuity of HIV/TB services, health system resilience, and hazard-specific preparedness.

**Funding:** National Institute for Health and Care Research (NIHR).

**Research in context:** *Evidence before this study:* We searched PubMed and Google Scholar using terms combining “health vulnerability”, “climate and health”, “composite indicator”, “district level”, “South Africa”, “HIV mortality”, and “TB mortality”, and reviewed reference lists in relevant IPCC and WHO reports on climate and health vulnerability assessments. No prior district-level vulnerability index for South Africa incorporating HIV/TB health outcomes and health systems was identified.

*Added value of this study:* We developed the first district-level HIV/TB Vulnerability Index for South Africa, integrating 27 indicators of HIV and TB outcomes, health system capacity, and socio-economic sensitivity and adaptation. We also developed a companion HIV/TB Vulnerability and Hazard Index (HVHI) which combines vulnerability with exposure to climate and weather hazards to identify districts facing the greatest compound risks. The analysis results revealed marked geographical disparities: the highest vulnerability was concentrated in KwaZulu-Natal, Eastern Cape, and Northern Cape, whereas districts in Gauteng and Western Cape exhibited greater adaptive capacity. The Hazard Index revealed a distinct east-west gradient with western districts experiencing greater heat stress and eastern districts facing higher flood and heavy rainfall hazards.

*Implications of all the available evidence:* Districts where climate and weather hazards coincide with high HIV/TB burden and constrained health system capacity face compounded risks, and should be prioritised for climate adaptation. Strengthening health system resilience including workforce capacity, hospital bed availability, primary health care access, and continuity of HIV/TB services, should be complemented by hazard-specific preparedness with heat-health planning prioritised in western districts and flood resilience in eastern districts. Investments in socio-economic enablers, including access to piped water, paved roads, and information are also essential. These findings provide an evidence base for geographically targeted climate-health adaptation planning in South Africa.

## Introduction

The health impacts of climate change are increasingly obvious, especially in low- and middle-income countries already burdened by communicable diseases and under-resourced health systems. ^1^ In sub-Saharan Africa, intensifying droughts, floods, and heat from climate change are disrupting health services, worsening infectious and nutritional diseases, increasing heat-related deaths and mental health burdens, and heightening food insecurity and vulnerable population displacement. ^2,3^

The South African Weather Service reports a clear shift toward more extreme conditions with rising maximum temperatures and more intense rainfall events. ^4^ The mean annual surface temperature has risen by 1.2°C, and the six hottest years on record have occurred within the past decade. Climate change has amplified the frequency and severity of heatwaves, droughts, and extreme rainfall, including the 2022 Durban floods. ^5^ These hazards heighten risks of food insecurity, malnutrition, and climate-sensitive infectious diseases. These risks are particularly pronounced for people living with HIV and Tuberculosis (TB), who face not only increased health risks during extreme weather events (EWEs) but also disruptions to prevention, diagnosis, treatment and continuity of care, leading to poorer outcomes. ^6–9^ Given the substantial burden of HIV/TB on South Africa’s health system and their major contribution to morbidity and mortality, climate variability and EWEs present a compounded public health challenge that threatens both population health and health system resilience.^10,11^

National climate adaptation frameworks rarely integrate health sector needs or address these specific vulnerabilities. ^12^ The capacity of health facilities to support community resilience during EWEs varies depending on location, infrastructure, staffing, and pre-existing community vulnerabilities. South Africa’s district health system (DHS), established under the National Health Act (2003), provides the decentralized framework for primary healthcare delivery. ^13^ Despite a uniform legislative framework, district-level performance is highly variable. Universal Health Coverage Service Coverage Index (UHC SCI) scores differ significantly among districts, and while maternal and child health services have improved, rural and under-resourced districts continue to lag in basic services provision. ^14–17^

Composite vulnerability indices are increasingly used to identify populations and health systems at greatest risk from climate change and to support adaptation planning. ^18–22^ Existing climate vulnerability assessments have generally focused on socio-economic vulnerability and settlement resilience, while climate-health vulnerability on broad population health and health service risks, or specific hazards such as heat or flooding, with relatively few incorporating disease-specific outcomes together with measured health system adaptive capacity. In South Africa, resources such as the Green Book, the South African Risk and Vulnerability Atlas, and the Health Risk and Vulnerability Assessment Tool for the National Department of Health provide valuable evidence on climate hazards, settlement vulnerability, and estimated health service risks. ^23–25^ However, none explicitly integrates observed HIV and TB burden, routinely measured health system performance, socio-economic determinants, and climate hazards within a single district-level framework.

Given that HIV and TB remain the leading causes of morbidity and mortality in South Africa and are particularly susceptible to disruption by climate variability and extreme weather events, we developed a disease-specific vulnerability framework centred on HIV and TB. This study therefore aimed to develop and apply a district-level HIV/TB Vulnerability Index, grounded in the IPCC conceptualisation of vulnerability, together with a complementary HIV/TB Vulnerability and Hazard Index to identify districts where high HIV/TB vulnerability coincides with high climate and weather hazard potential. By integrating epidemiological, health system, socio-economic, and hazard indicators, our analysis provides a spatially explicit assessment of climate and HIV/TB vulnerability at the district level, the primary operational unit for service delivery. The resulting framework is intended to support policymakers and health planners in prioritising adaptation investments, targeting interventions, and establishing a baseline for monitoring changes in climate-related health vulnerability over time.

This study, therefore, aimed to a) assess and map district-level exposure to climate and weather hazards, including floods, storms, heatwaves and droughts, alongside vulnerabilities related to HIV and TB burden and health system capacity, and b) develop a composite HIV/TB Vulnerability Index and a compound vulnerability-hazard index to identify districts where high HIV/TB vulnerability coincides with high hazard exposure. By integrating epidemiological, health system, socioeconomic and hazard indicators, our analysis provides a spatially explicit assessment of district-level climate-related vulnerability. The resulting framework is intended to support policymakers and health planners in prioritising adaptation investments, targeting interventions, and establishing a baseline for monitoring changes in climate-related health vulnerability over time.

## Methods

To quantify district-level vulnerability, we apply a composite indicator methodology which captures complex or multidimensional phenomena by combining multiple individual indicators into a single summary measure. Composite indicators are widely used to assess concepts such as well-being, happiness, vulnerability, livelihoods, sustainability or innovation. ^26^

The construction of a composite indicator follows a structured and well-established methodological framework that has been described extensively in handbooks and applied across numerous studies. Key concepts include: developing a conceptual framework, identifying and evaluating data sources, selecting and processing indicators, normalising indicator values, weighting and aggregating indicators within subdomains; constructing intermediate and overall composite scores; visualising the results; and conducting validation and sensitivity analyses to assess the robustness of the index. ^26–28^

To establish the theoretical framework we adopted the most recent definition of vulnerability proposed by the IPCC which conceptualises vulnerability as a multidimensional construct comprising two core components: *sensitivity and adaptative capacity*. Each component encompasses multiple dimensions that reflect the characteristics the framework seeks to capture within the composite measure including health, socio-economic conditions and, the broader environmental context.

In this analysis, vulnerability was conceptualised across three dimensions: i) HIV and TB health outcomes, ii) health system capacity, and iii) socio-economic conditions encompassing demographic and socio-economic indicators. ^29^ Grouping indicators into these dimensions provides a clearer understanding of the underlying drivers of the composite index and enhances interpretability. ^21,26^

Indicators were assigned to either sensitivity or adaptive capacity according to criteria consistent with the IPCC vulnerability framework. Indicators were classified as sensitivity if they reflected the current state or outcomes that determine the degree of susceptibility to harm from a given exposure. Indicators were classified as adaptive capacity if they represented the resources, assets, or institutional capacities that enable individuals, communities, or health systems to anticipate, respond to, and recover from future exposures.

This classification was applied at the level of individual indicators. Consequently, although the socio-economic dimension comprises indicators assigned to both sensitivity and adaptive capacity, each indicator was assigned exclusively to one component and not both.

Within the HIV/TB dimension, all indicators represent realised health outcomes (mortality, prevalence, years of life lost) and were therefore classified as sensitivity. Indicators representing the capacity to prevent, manage or respond to HIV and TB are captured within the health systems dimension, where measures such as antiretroviral therapy (ART) retention, health facility density, hospital bed density and healthcare workforce density were classified as adaptive capacity.

Within the socio-economic dimension, indicators reflecting existing socioeconomic disadvantage (e.g., poverty, unemployment, dependency ratio) were classified as sensitivity, whereas indicators representing resources and assets that enable individuals and communities to anticipate, respond to and recover from adverse events (e.g., access to paved roads, piped water, higher education attainment, and access to information) were classified as adaptive capacity. The complete list of indicators and the rationale for their classification are provided in Table 1.

**Table 1.**
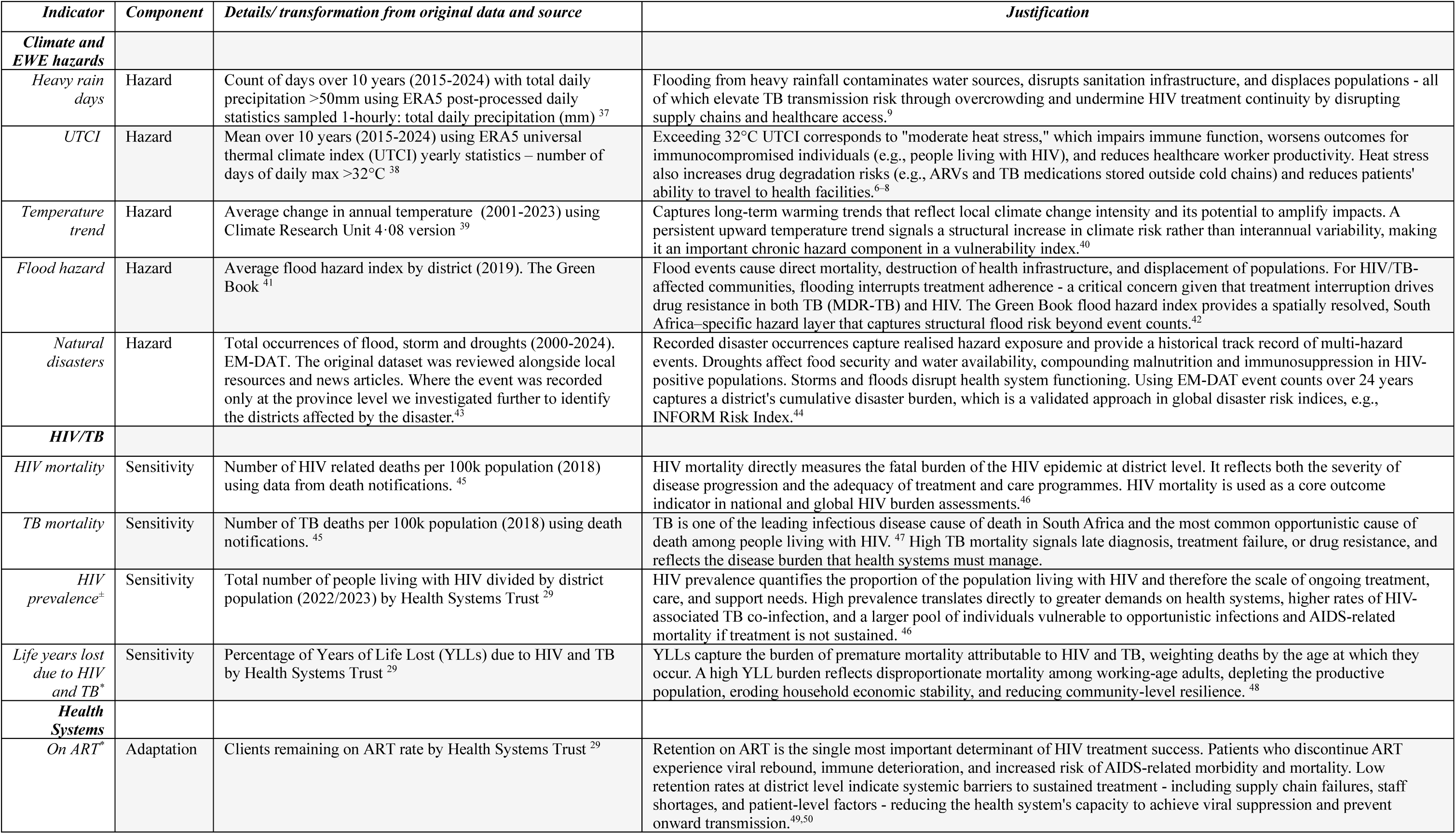
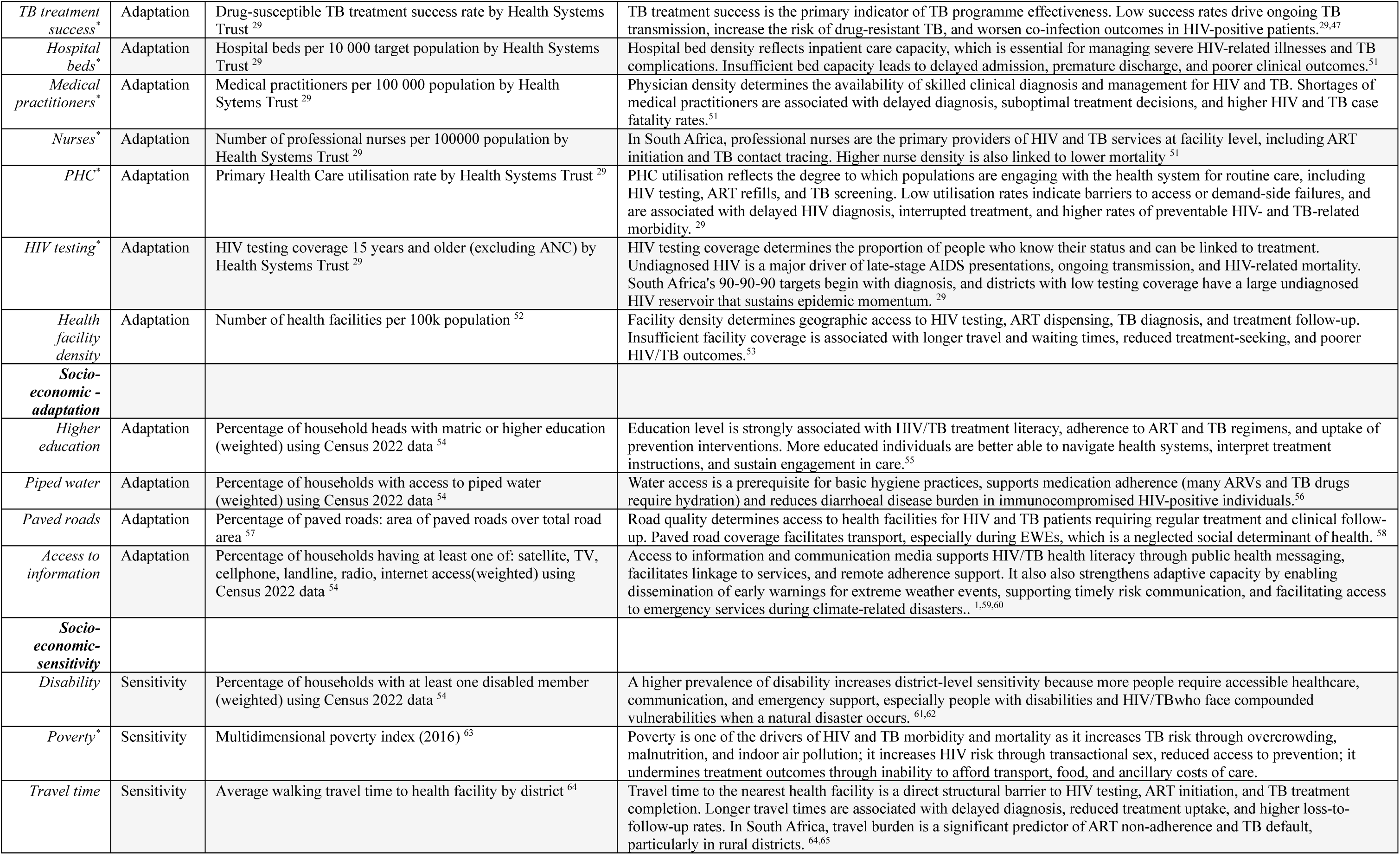
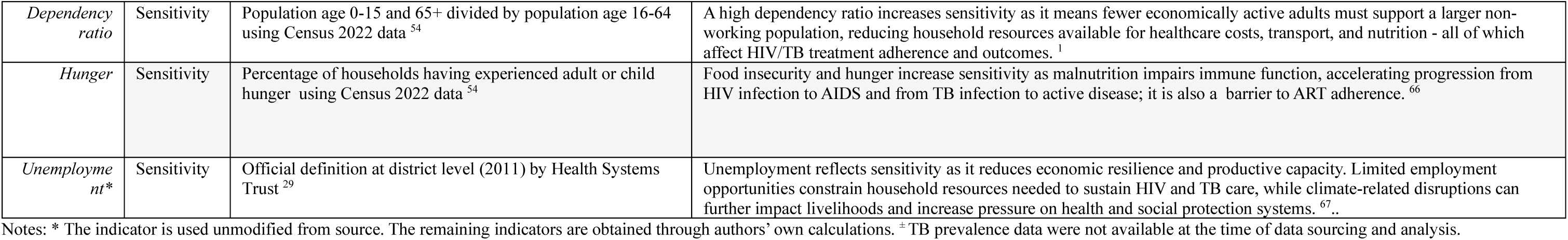
Indicators: data source, transformation and justification.

Indicator selection followed an iterative, theory-driven process. An initial pool of 80 candidate indicators aligned with the conceptual framework was identified from multiple data sources, drawing on previous research, expert input and frameworks developed by international organisations.^1,20,22,26,30^ Candidate indicators were mapped to the conceptual framework (Figure 1) to ensure alignment with the underlying theoretical constructs of sensitivity and adaptive capacity. Although alternative conceptual classifications are plausible for some indicators, particularly socio-economic infrastructure variables, each indicator was assigned according to a predefined theoretical criterion derived from the updated IPCC framework to ensure conceptual consistency across the index.

**Figure 1.**
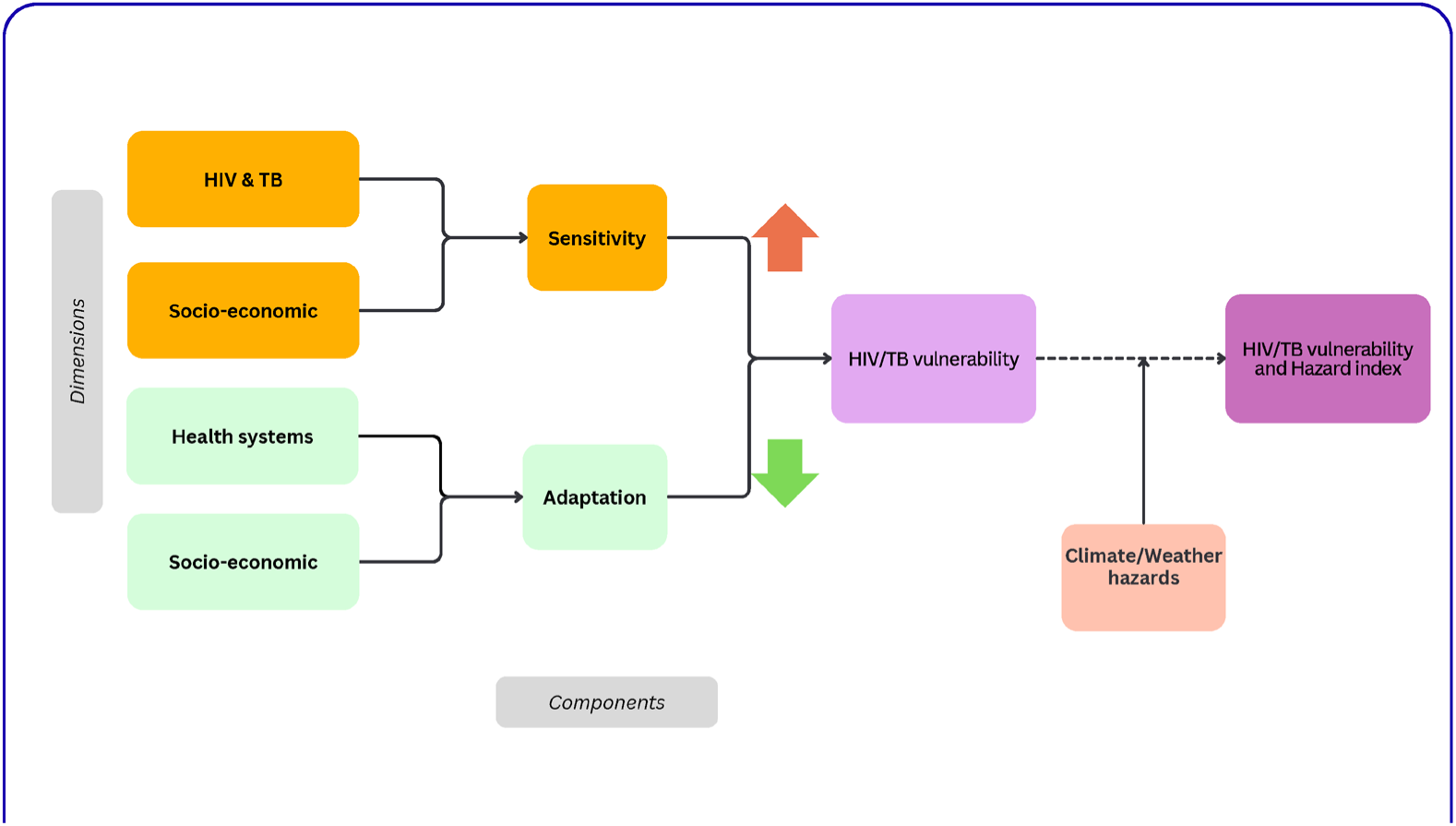
HIV/TB health vulnerability framework.

From this initial pool, 27 indicators were retained for the final index. Selection was guided by three criteria: (1) conceptual relevance to the HIV/TB mortality and health systems vulnerability framework; (2) availability and quality of district level data ; and (3) statistical non-redundancy. To assess redundancy, pairwise Pearson correlation coefficients were examined within each component, and indicators with an absolute corelation coefficient |r| > 0.80 were considered for exclusion when they captured substantively overlapping constructs. For example, walking time and motorised travel time to the nearest health facility were highly correlated; therefore only walking time was retained.

Indicators that showed little or no association with any other indicator within the same dimension were also considered for exclusion, as they were unlikely to contribute meaningfully to the latent construct represented by that dimension. Land-use and land-cover indicators were similarly evaluated but were not included because they were considered insufficiently informative at the district scale - although they may be appropriate for composite indices at finer spatial resolutions.

Final decisions on indicator inclusion balanced statistical considerations with conceptual importance, ensuring that each retained indicator contributed distinct non-overlapping information to its respective component while preserving the theoretical integrity of the composite index. The correlation structure of the final 27 indicators by dimension shows low redundancy with only three correlation coefficients above |0.8|: access to piped water and poverty index (-0.88), access to piped water and dependency ratio (-0.86), and dependency ratio and poverty (0.84) (supplementary material page 8). We decided to include these indicators as they are considered to reflect different aspects of socio-economic conditions.

To construct the composite indices, we adopted a hierarchical, deductive design and conducted sensitivity analyses to assess the influence of methodological design choices on district vulnerability rankings (supplementary material pages 9-12). ^31^

All selected indicators were first normalised using min-max scaling which transforms each indicator to a value ranging from zero (lowest observed district value) to one (highest observed district value). The normalised indicators were then grouped into the four dimensions defined by the conceptual framework - HIV/TB outcomes, health system capacity, socio-economic sensitivity, and socio-economic adaptative capacity - and aggregated using equal weighting to generate intermediate dimension scores 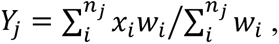 where *j* = 1, 2, 3 denotes the dimension, *i* = 1, …, *n_j_* denotes the indicator *x_j_* within dimension *j* for each district.

Equal weighting was selected because it is transparent, intuitive and readily interpretable when communicating findings to policymakers and other stakeholders. It is also the most commonly adopted approach in composite indicator construction when robust empirical or context specific evidence for differential weighting is lacking. ^20,26,32^

The same equal-weighting and aggregation procedure was subsequently applied to derive composite scores for the two overarching components of vulnerability: sensitivity and adaptative capacity. This hierarchical approach preserves the conceptual structure of the vulnerability framework, generates interpretable scores at both the dimension and component levels, and prevents dimensions containing larger numbers of indicators from disproportionately influencing the overall index.

Finally, the HIV/TB Vulnerability Index (HVI) was calculated using an additive model defined as:

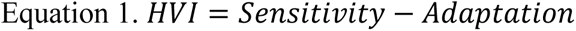

This framework is consistent with the most recent Intergovernmental Panel on Climate Change definition of vulnerability which conceptualises vulnerability independent of exposure.^1^ In addition, we developed a Climate and Weather Hazard index, that can be combined with the HVI to derive a composite HIV/TB Vulnerability and Hazard Index (HVHI). The Climate and Weather Hazard Index includes complementary indicators that represent different aspects of climate hazards. Specifically, the Universal Thermal Climate Index (UTCI) captures the frequency of heat stress days (>32°C) and therefore reflects exposure to extreme heat conditions, whereas the temperature trend indicator characterises long-term climatic warming. In addition, the index incorporates the historical occurrence of floods, storms, and droughts (derived from EM-DAT), together with flood hazard and heavy rainfall indicators, to capture other dimensions of climate and weather hazards.

The combined HVHI identifies districts where high underlying vulnerability coincides with high climate and weather hazards thereby highlighting priority areas for adaptation and resilience planning. As the HVHI integrates both vulnerability and hazard, it is intended as an operational planning tool and should not be interpreted as a measure of vulnerability itself.

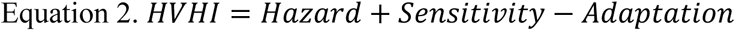

For ease of interpretation and presentation, all intermediate and final composite indices were rescaled to a 0-1 range using min-max normalisation. This transformation preserves the relative ordering of districts and does not affect the vulnerability rankings.

The final composite indicators were constructed using an additive aggregation model, whereby greater sensitivity increases vulnerability while greater adaptive capacity reduces it. The additive structure was selected because it is transparent, readily interpretable by policymakers and other stakeholders, and consistent with established guidance for the development of composite indicators. It should therefore be interpreted as a pragmatic aggregation rule for summarising multiple dimensions of vulnerability rather than as a causal model of the underlying relationships between the individual components.

The formulation should not be interpreted as implying that the underlying relationships between sensitivity, adaptive capacity and vulnerability are strictly linear. Rather the additive model represents a pragmatic and widely accepted aggregation approach in the absence of empirical evidence demonstrating that alternative functional forms provide a more appropriate representation of vulnerability. ^31^

Finally, the robustness of the composite indicator was evaluated through sensitivity analyses to assess the extent to which methodological choices influenced the identification and ranking of vulnerable districts. ^31^. Specifically, we examined the effects of alternative index structures, comparing the current theory driven (deductive) aggregation approach with a data-driven (inductive) approach based on principal component analysis (PCA) as well as alternative normalisation and weighting methods (supplementary material pages 9-12).

Overall, district vulnerability rankings were robust to these plausible methodological alternatives. Different normalisation procedures had negligible effects on district rankings, while PCA-derived weighting schemes resulted in only modest changes. Importantly, the districts consistently identified as the most vulnerable remained largely unchanged across all specifications, indicating that the principal spatial patterns reported were not artefacts of the chosen normalisation, weighting or aggregation strategy.

We also assessed the influence of individual indicators on district rankings. These analyses demonstrated that no single indicator exerted undue influence on the overall index, confirming that the final indicator set is both robust and non-redundant (supplementary material pages 11-12).

### Data

South Africa’s 52 districts represent the second level of administrative division below the nine provinces. Of these, 44 are district municipalities while the remaining eight correspond to the largest urban metropolitan municipalities. Districts differ markedly in area, population size, governance structures and level of development. Table 1 presents the final set of indicators, including definitions, data sources, justification and any transformations applied to the original datasets, while summary statistics of the indicators can be found in page 2 of supplementary material. Where more recent district-level estimates were unavailable, the latest reliable data were retained for indicators representing relatively stable structural characteristics or conceptually essential components of vulnerability.

Data processing and analysis was done in R version 4.4.1.^33–37^

## Results

### Climate and extreme weather hazards

To characterise climate hazard variability across districts, we included the following indicators: number of natural disasters (2001-2023), flood hazard index (2019), mean temperature trend (2001-2023), number of days with heavy rain (>50 mm), and number of annual UTCI days exceeding 32°C daily max (Table 1). The temperature indicators capture both chronic thermal exposure relevant to heat-related illness, and long-term warming trends that reflect local climate change intensity and its potential to amplify impacts. The flood hazard index combines historical flood occurrence, hydrological modelling, and topographic data to provide a spatially explicit measure of district-level flood risk.^38^ Western districts are more affected by high temperatures, while those in the east by flood and heavy rainfall hazards (Figure 2). Districts in the Eastern Cape province have experienced the highest number of recorded disasters (storms, floods and droughts) in the last 25 years with O.R. Tambo district recording the highest number (10). Extreme heat conditions affect mostly the western part of the country (Z.F. Mgcawu district records 184 UTCI). Heavy rainfall (>50 mm) impacts districts in the east coast with eThekwini district reaching 12·3 days with heavy rainfall over 10 years (page 2 in supplementary material). Figure 2 presents the spatial patterns of the composite Climate and EWEs hazard score showing that the combined weather hazard is higher among districts in the east.

**Figure S1:**
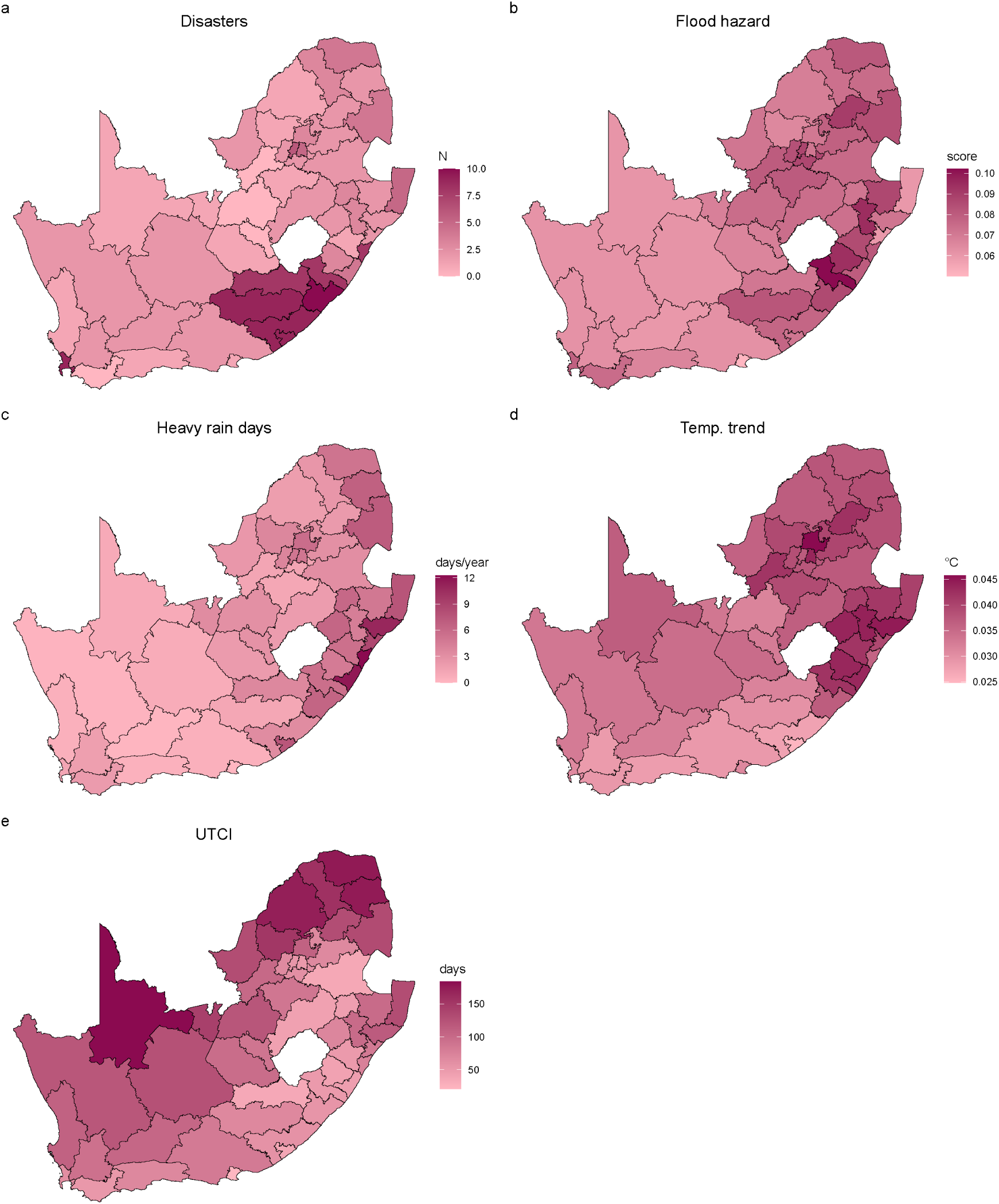
Climate and weather hazards.

### HIV and TB health outcomes

This dimension includes HIV prevalence, HIV and TB-related mortality and the percentage of years of life lost (YLL) attributable to these diseases (supplementary material page 2). HIV mortality averages at 44·8 deaths per 100,000 population, ranging from 3 in Ekurhuleni to 94 in John Taolo Gaetsewe. TB mortality was lowest in the City of Johannesburg (20 per 100,000) and highest in Chris Hani District (101 per 100,000). HIV and TB mortality spatial patterns were not necessarily linked to disease burden which is highest in KwaZulu-Natal (supplementary material page 4). The percentage of YLL due to HIV and TB followed more closely the prevalence patterns suggesting a high impact of preventable deaths among younger populations in the same areas.

### Health system capacity

Most health system indicators were obtained from the most recent District Health Barometer (2022/2023) compiled by the Health Systems Trust (HST).(Ndlovu N & Padarath A, 2024) These included hospital beds and medical practitioners per target population, primary health care (PHC) utilisation rate, TB treatment success rate, and proportion of people living with HIV that remained on treatment. These indicators were interpreted as ‘adaptive capacity’ as they captured health system responsiveness, treatment coverage, surge capacity and routine service delivery (Table 1).

Health system capacity differed substantially between indicators and across districts. With respect to capacity (facilities, beds, medical staff), rural and large area districts were more affected than urban districts. A north-south divide was evident when looking at the number of health facilities per 100,000 population, with the north having less facilities than the south ( supplementary material page 5). Health personnel was limited in rural and remote areas where long distances made access to hospitals, emergency care, and recruiting and retaining skilled health professionals difficult. However, several rural districts performed well in terms of treatment coverage and routine service delivery, for example, PHC utilisation was highest in uMkhanyakude district (3·2 visits per person per year) - noting that the national target is 3·5; high ART uptake rate and TB treatment success was also linked to low density districts (supplementary material page 5).^30^ Moreover, four out of the five districts scoring high in health systems adaptation were rural districts: the top three performing districts in health systems adaptation were predominantly rural.

### Socio-economic

The socio-economic dimension encompasses demographic, socio-economic and infrastructural factors that influence a population’s sensitivity and capacity to adapt to climate and health-related shocks consistent with established vulnerability frameworks.^19,39^ Indicators such as low educational attainment, high dependency ratios, poverty and limited access to piped water were conceptualised as increasing sensitivity by constraining adaptive knowledge, placing greater economic strain on households, and exacerbating health risks. In contrast, access to communication media, paved roads, piped water, higher educational attainment and shorter travel times to health facilities were considered indicators of adaptive capacity because they facilitate access to services, information, and social and economic resources that support effective responses to adverse events.

Adaptive capacity was highest in urban and metropolitan districts, where access to piped water and higher education was greater and travel times to the nearest health facility were shorter. Conversely, many rural districts in the Eastern Cape, North-West and KwaZulu-Natal exhibited high socioeconomic sensitivity reflecting elevated levels of poverty, food insecurity, dependency and disability. These findings highlight marked geographic inequalities in the socioeconomic determinants of vulnerability across South Africa (supplementary material pages 6-7).

### HIV/TB Vulnerability Index

According to the composite HVI (Figure 3 and 4), the ten most vulnerable districts were John Taolo Gaetsewe (Northern Cape), O.R. Tambo (Eastern Cape), Z.F. Mgcawu (Northern Cape), Dr Ruth Segomotsi Mopati (North-West), Alfred Nzo (Eastern Cape), Harry Gwala, uMkhanyakude (KwaZulu-Natal), Pixley Ka Seme (Northern Cape), uThukela and Ugu (KwaZulu-Natal). In contrast, the least vulnerable districts were either a metro district or located in Gauteng and the Western Cape provinces: City of Tshwane, City of Johannesburg, Ekurhuleni, City of Cape Town and Overberg.

**Figure 3:**
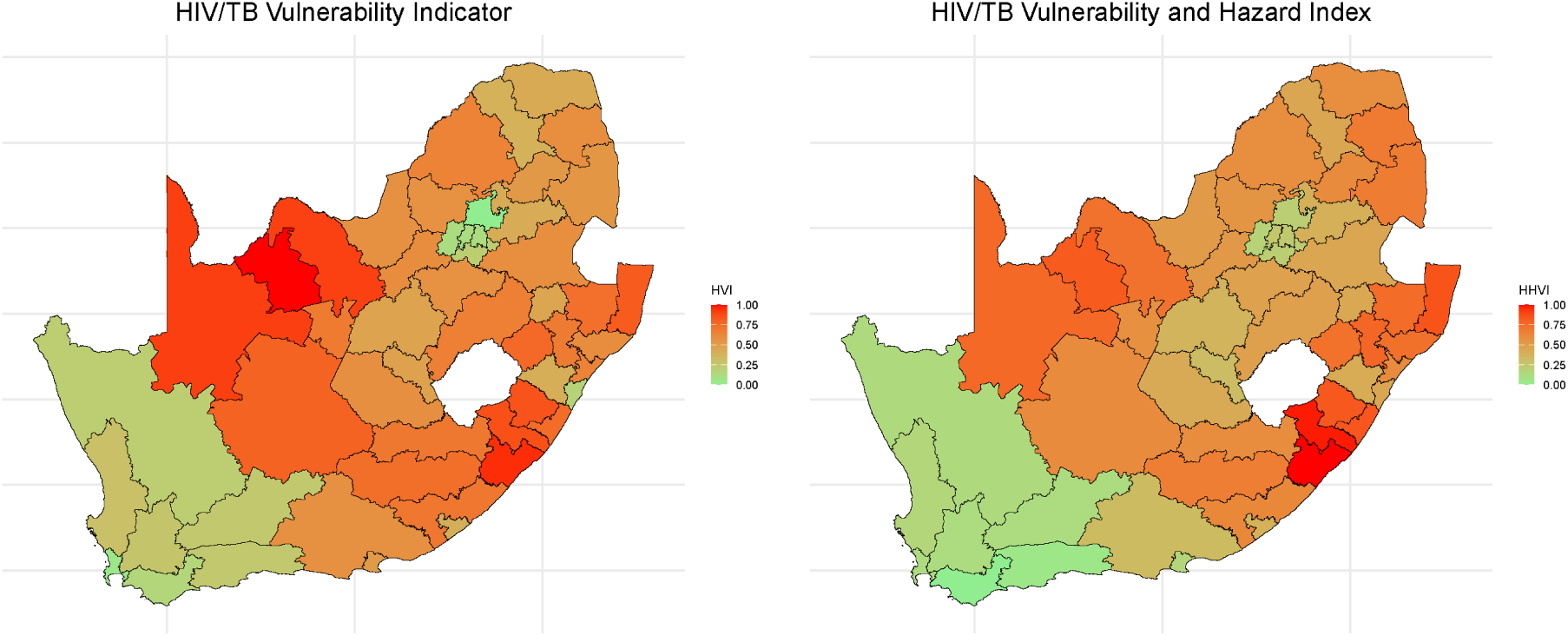
HIV/TB Vulnerability versus HIV/TB Vulnerability and Hazard Index. Shading indicates vulnerability level, ranging from red (highest vulnerability) to green (lowest vulnerability).

**Figure 4:**
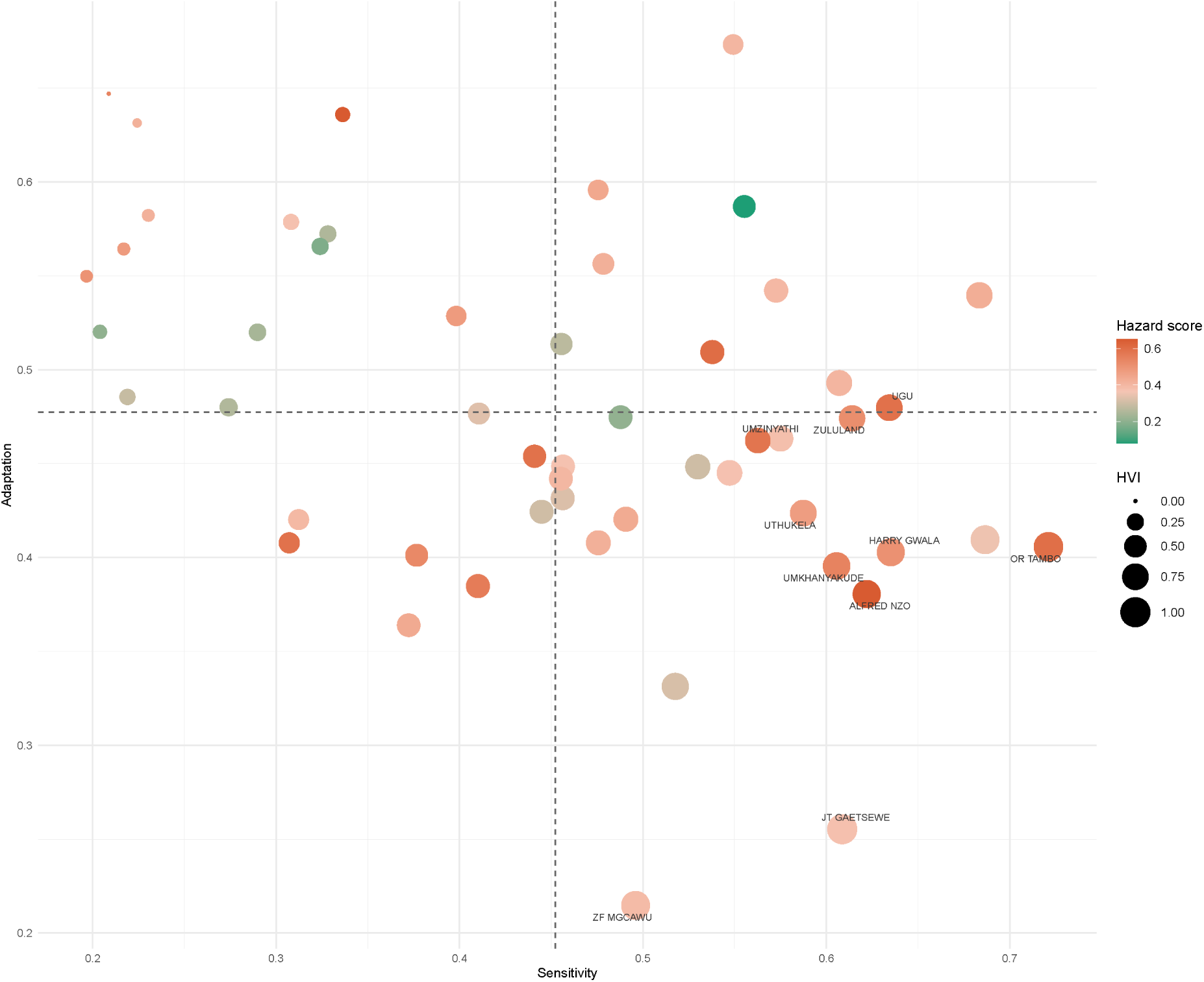
Component contributions to the HIV/TB Vulnerability Index (HVI) by district and province. Districts are positioned by sensitivity (x-axis) and adaptive capacity (y-axis). Dashed lines represent the average scores. Quadrants illustrate relative vulnerability profiles using HHVI: upper-left = high adaptive capacity/low sensitivity (low vulnerability); lower-left = below-average sensitivity and adaptation (moderate vulnerability); lower-right = high sensitivity/low adaptive capacity (high vulnerability). Top 10 most vulnerable districts are labelled.

After incorporating the Climate and Weather Hazard Index to derive the HIV/TB Vulnerability and Hazard Index (HVHI), eight out of the ten highest scoring districts are along South Africa’s east coast, four in KwaZulu-Natal and two in the Eastern Cape. These districts are characterised by the convergence of high climate hazards, substantial socioeconomic disadvantage and high HIV/TB burden. The remaining two high scoring districts are in the Northern Cape province where particular hazards intersect with low health systems adaptive capacity.

Overall, eight of the ten districts ranked most vulnerable by the HVHI were also among the top ten according to the HVI, indicating that underlying vulnerability was the principal driver of overall risk. However, two additional KwaZulu-Natal districts, uMzinyathi and Zululand entered the top ten after the inclusion of climate hazards, demonstrating that elevated hazard exposure can substantially alter adaptation priorities despite comparatively lower underlying vulnerability.

Figure 4 illustrates the relationship between sensitivity and adaptive capacity for each district with climate hazard overlaid to illustrate how they contribute to overall vulnerability. Districts in the upper left quadrant exhibit relatively low sensitivity and high adaptive capacity, resulting in low overall HIV/TB vulnerability. Districts in the lower left quadrant have lower average scores for both sensitivity and adaptive capacity, yielding intermediate levels of vulnerability. In contrast, districts in the lower right quadrant combine above-average sensitivity with below-average adaptive capacity, placing them amongst the most vulnerable.

Metropolitan districts in the Western Cape and Gauteng cluster in the upper left quadrant, reflecting low sensitivity alongside high adaptive capacity. Their stronger health systems, more developed infrastructure, and more favourable socio-economic conditions contribute to lower overall vulnerability. Consequently, these districts maintain low scores, even after climate hazards are incorporated, suggesting greater resilience to the health impacts of climate and weather related events.

To better understand the drivers of district vulnerability, we examined the relative contributions of the individual components and dimensions by identifying the highest scoring element for each district. Table 2 shows the dominant component and dimension among the top most vulnerable district rankings ordered by their overall HIV/TB Vulnerability and Hazard score. Eight of ten most HVI-vulnerable districts are also in the top ten scoring in HVHI. Dominance in an additive model is determined by the highest score. When looking at the HVHI, the dominant component appears to be sensitivity in eight districts and hazard score in two districts. Within sensitivity, HIV/TB dimension scores higher than socio-economic dimension for eight districts highlighting that their vulnerability is greatly driven by unfavourable HIV/TB outcomes. Similarly, the dimension that drives adaptation is health systems adaptive capacity. Among the ten highest ranked districts, sensitivity was the dominant component in eight whereas climate hazard component was dominant in the remaining two.

**Table 2.** Dominant component and dimension among the 10 districts with the highest HIV/TB vulnerability and Hazard score.

| Prov. | District | HVI | HVI rank | HVHI | HVHI rank | Dominant Component | Dominant sensitivity dimension | Dominant adaptation dimension |
| --- | --- | --- | --- | --- | --- | --- | --- | --- |
| EC | OR TAMBO | 0.95 | 2 | 1 | 1 | Sensitivity | HIV/TB. | Health systems |
| EC | ALFRED NZO | 0.86 | 5 | 0.98 | 2 | Hazard | Socio-economic | Health systems |
| KZN | UMKHANYA KUDE | 0.82 | 7 | 0.85 | 3 | Sensitivity | HIV/TB. | Health systems |
| KZN | UGU | 0.75 | 10 | 0.84 | 4 | Sensitivity | HIV/TB. | Health systems |
| KZN | HARRY GWALA | 0.85 | 6 | 0.83 | 5 | Sensitivity | HIV/TB. | Health systems |
| NC | JOHN TAOLO GAETSEWE | 1 | 1 | 0.82 | 6 | Sensitivity | HIV/TB. | Socio-economic |
| KZN | UMZINYATH I | 0.68 | 16 | 0.78 | 7 | Hazard | Socio-economic | Health systems |
| NC | ZF MGCAWU | 0.91 | 3 | 0.76 | 8 | Sensitivity | HIV/TB. | Health systems |
| KZN | ZULULAND | 0.73 | 12 | 0.75 | 9 | Sensitivity | HIV/TB. | Health systems |
| KZN | UTHUKELA | 0.76 | 9 | 0.73 | 10 | Sensitivity | HIV/TB. | Health systems |
| KZN | UTHUKELA | 0.76 | 9 | 0.73 | 10 | Sensitivity | HIV/TB. | Health systems |

Although adaptive capacity partially mitigated vulnerability in these districts, it was insufficient to offset the high levels of sensitivity. In particular, the socioeconomic adaptive capacity dimension consistently recorded relatively low scores among the ten most vulnerable districts, suggesting limited socio-economic resources to buffer the HIV/TB sensitivity (supplementary material pages 12-13).

Within each dimension, we observed similar patterns across individual indicators with the exception of the two North-West districts that showed a hazard profile driven almost entirely by extreme heat stress (UTCI), rather than disasters, floods, or rainfall (supplementary material pages 13-14). The sensitivity of Z.F. Mgcawu district is dominated by extraordinary long travel time to the nearest health facility showing the challenges in accessing health care in large and sparsely populated districts with poor infrastructure and road systems. Additionally, Z.F. Mgcawu district is characterised by extremely low adaptive capacity in its health system which failed to offset it sensitivity and lower overall vulnerability. uMkhanyakude district in KwaZulu-Natal province was affected by both high temperatures and relatively strong rainfall and was deemed vulnerable due to the high socio-economic sensitivity and equally low socio-economic adaptive capacity. It presented notably strong health-system service delivery, especially on ART coverage, PHC utilisation, and TB treatment success rate, although lagging on capacity (facility and medical practitioner density). It’s overall high vulnerability was also due to high HIV prevalence and mortality.

## Discussion

This study developed South Africa’s first district-level HIV/TB Vulnerability Index (HVI), integrating HIV and TB outcomes, health system capacity, and socio-economic conditions within an IPCC-grounded vulnerability framework. A companion HIV/TB Vulnerability and Hazard Index (HVHI) identified districts where high underlying vulnerability coincides with high climate and weather hazards. Vulnerability was concentrated in KwaZulu-Natal, the Eastern Cape, and the Northern Cape, while Gauteng and the Western Cape consistently showed lower vulnerability, reflecting stronger health systems, more favourable socio-economic conditions, and greater adaptive capacity.

Notably, vulnerability in most high-ranking districts was driven primarily by poor HIV/TB outcomes and not socio-economic disadvantage alone: HIV/TB burden was the dominant sensitivity dimension in eight of the ten most vulnerable districts. Health system adaptive capacity also generally contributed more than socio-economic adaptive capacity in the overall adaptation component. This suggests that strengthening health systems is an immediate, actionable lever for reducing vulnerability, while broader socio-economic gains remain essential for long-term resilience.

The rural-urban pattern is more nuanced than health system capacity alone would suggest. Metropolitan districts benefit from greater infrastructure, workforce density, and more favourable socio-economic conditions, yet several rural districts in KwaZulu-Natal and the Eastern Cape achieve comparatively strong HIV/TB service delivery despite limited resources - for example, primary healthcare utilisation is highest in uMkhanyakude, and several rural districts perform well on ART retention and TB treatment success. This suggests vulnerability in these settings stems mainly from high underlying HIV/TB burden, socio-economic disadvantage, and lack of capacity instead of poor healthcare delivery.

These patterns are further highlighted by the intersection with climate hazards: high hazard scores overlap with high vulnerability in eight out of the ten most vulnerable districts. The hazard index shows a distinct east-west split western districts face greater chronic heat stress, while eastern coastal districts face higher flood risk and heavy rainfall. This is consistent with prior climate risk assessments of South Africa. Eastern districts combine high hazard with high HIV/TB burden and socio-economic disadvantage; western districts face greater heat stress alongside weaker health systems and similarly adverse socio-economic conditions. The HVHI adds further value by identifying districts where climate hazard substantially shifts overall priority: in Alfred Nzo and uMzinyathi, hazard became the dominant contributor despite lower underlying vulnerability, and Zululand rose into the top ten once hazard was included, reflecting the benefit of integrating hazard and vulnerability for adaptation planning.

Finally, the socio-economic dimension shows that access to piped water, paved roads, information, and higher education are not merely markers of development but key determinants of district-level adaptive capacity. The Eastern Cape and North West consistently recorded the lowest adaptive capacity scores on these indicators, underscoring structural disadvantages that constrain resilience. Strengthening these socio-economic assets should therefore be treated as integral to climate adaptation for HIV/TB programmes, not a separate development objective.

Our findings are broadly consistent with previous South African vulnerability assessments, including the Green Book, which identified rural, resource-constrained districts as disproportionately vulnerable to climate-related risks. ^23–25^ Internationally, climate-health vulnerability indices have primarily been developed to identify populations at risk from climate change by combining measures of climate hazards with socio-economic, demographic, environmental, or health system characteristics, most commonly for heat-related health risks, flooding, or general population health.^18,20,21,40^ Existing South African frameworks similarly focus on climate hazards, settlement resilience, socio-economic vulnerability, or health service risks, rather than HIV and TB. ^23,25^ Our index extends this literature by integrating HIV and TB outcomes, health system adaptive capacity, socio-economic conditions, and climate hazards within a single district-level framework. By placing HIV and TB at the centre of the vulnerability assessment and using routinely collected district-level health indicators, the index provides a practical basis for prioritising adaptation investments in settings where climate hazards intersect with a high burden of infectious diseases.

Several policy implications emerge from these findings. First, adaptation investments should prioritise districts in KwaZulu-Natal, the Eastern Cape, and the Northern Cape, where high HIV/TB burden coincides with limited adaptive capacity. Strengthening the health workforce, primary healthcare, and continuity of HIV and TB services should be central to these efforts. Second, adaptation strategies should be hazard-specific, with heat-health action plans in western districts and flood preparedness, transport resilience, and supply chain continuity in the east. Third, socio-economic investments, including access to safe water, transport infrastructure, education, and information, should be recognised as core components of health resilience rather than separate development priorities. The index identifies districts where multiple structural drivers of vulnerability converge, but it should not be interpreted as evidence that improving any single indicator will directly reduce HIV or TB outcomes. Instead, the findings support integrated, multi-sectoral adaptation strategies that address multiple dimensions of vulnerability simultaneously.

This study has several limitations. First, the index is based on the most recent district-level data available, with indicators spanning 2011-2024, although the oldest indicators represent relatively stable structural characteristics. As newer data become available, the index should be updated. In addition, district-level HIV/TB mortality estimates are limited to the latest available mortality data, although these diseases remain the leading causes of death in South Africa. ^10^ Second, consistent with the updated IPCC framework, the HVI characterises vulnerability rather than overall climate risk because suitable district-level exposure metrics are not yet available. ^41^ The companion Climate and Weather Hazard Index uses EM-DAT disaster occurrence data, which may underestimate smaller, locally significant events and is influenced by reporting completeness. However, this limitation is partly mitigated by combining EM-DAT with independently derived indicators of heat stress, long-term temperature trends, flood hazard, and heavy rainfall. The original EM-DAT dataset was reviewed alongside local resources and news articles. Where the event was recorded only at the province level we investigated further to identify the districts affected by the disaster. Moreover, the HVI was intentionally designed as a hazard-independent measure of vulnerability according to the most recent IPCC definition, and therefore does not distinguish vulnerability according to specific hazard types, such as heatwaves, floods, or droughts. Future research should extend this framework by incorporating spatially resolved exposure metrics and developing hazard-specific risk indices that capture the pathways through which different climate hazards affect HIV/TB health. Finally, no accepted external gold standard currently exists for validating multidimensional climate-health vulnerability indices. Although extensive sensitivity analyses demonstrated that district rankings were robust to alternative normalisation, weighting, and indicator selection methods, future studies should evaluate the validity of these indices by examining their ability to predict districts that experience disproportionately adverse HIV/TB health impacts during extreme weather events.

Despite these limitations, the HIV/TB Vulnerability Index and HIV/TB Vulnerability and Hazard Index provide transparent, reproducible tools for identifying where climate hazards, HIV/TB burden, and limited adaptive capacity converge. They offer a practical framework to inform district-level adaptation planning, prioritise climate-health investments, and support implementation of the World Health Organization Climate and Health Vulnerability and Adaptation Assessment framework and the United Nations Framework Convention on Climate Change National Adaptation Plan process. ^42,43^ Because the indices are based on publicly available data and can be readily updated as newer data become available, they provide a scalable approach for monitoring vulnerability over time and can be adapted to other high-burden diseases and settings. Although developed for South Africa, the framework is readily transferable to other low- and middle-income countries by selecting locally appropriate indicators while retaining the same conceptual structure. This provides a practical methodology for supporting climate adaptation planning in other settings where HIV, tuberculosis, or other climate-sensitive diseases place substantial demands on health systems

As climate hazards intensify, adaptation strategies should move beyond hazard mapping alone to address the underlying drivers of vulnerability. Integrating health system strengthening, continuity of HIV and TB services, and action on the socio-economic determinants of health will be essential for building climate-resilient health systems.

## Data Availability

All data produced in the present study are publicly available at cited sources or available upon reasonable request to the authors.

## Contributors

EG, MC and CI contributed to study conceptualisation and methods. NM and MC contributed to the data sourcing and processing. EG did the data processing, analysis, visualisation of figures and tables. EG, MC, SK, SB and CI contributed to the draft preparation. All authors had access to the data used in this analysis apart from the restricted access data (HIV and TB mortality), for which EG, CI and NM had full access.

## Acknowledgements

The ASTRA study was funded by the National Institute for Health and Care Research (NIHR) through an NIHR RIGHT award to the University of Sussex with reference NIHR204828. The views expressed are those of the author(s) and not necessarily those of NIHR or the Department of Health and Social Care. For the purpose of open access, the author has applied a Creative Commons Attribution (CC BY) licence to any Author Accepted Manuscript arising.

We would like to thank the South African Department of Health for their support of the ASTRA study. The Africa Health Research Institute, KwaZulu-Natal, which hosted the study receives strategic core grant from Wellcome (Ref: 227167/A/23/Z). Statistics South Africa for granting us access to mortality data.

## References

1 IPCC. Climate Change 2022 – Impacts, Adaptation and Vulnerability. Cambridge University Press, 2023 DOI:10.1017/9781009325844.

2 Chersich MF, Wright CY, Venter F, Rees H, Scorgie F, Erasmus B. Impacts of climate change on health and wellbeing in South Africa. International Journal of Environmental Research and Public Health 2018; 15. DOI:10.3390/ijerph15091884.

3 Wright CY, Kapwata T, Naidoo N, et al. Climate Change and Human Health in Africa in Relation to Opportunities to Strengthen Mitigating Potential and Adaptive Capacity: Strategies to Inform an African “Brains Trust”. Annals of Global Health 2024; 90. DOI:10.5334/aogh.4260.

4 South African Weather Service. Trends in Extreme Climate Indices in South Africa 2023. https://github.com/ARCCSS-.

5 Ziervogel G, Lennard C, Midgley G, et al. Climate change in South Africa: Risks and opportunities for climate-resilient development in the IPCC Sixth Assessment WGII Report. South African Journal of Science 2022; 118. DOI:10.17159/sajs.2022/14492.

6 Iwuji CC, Baisley K, Maoyi ML, et al. The Impact of Drought on HIV Care in Rural South Africa: An Interrupted Time Series Analysis. EcoHealth 2023; 20: 178–93.

7 Iwuji CC, McMichael C, Sibanda E, Orievulu KS, Austin K, Ebi KL. Extreme weather events and disruptions to HIV services: a systematic review. The Lancet HIV 2024; 11: e843–60.

8 Orievulu K, Ayeb-Karlsson S, Ngwenya N, et al. Economic, social and demographic impacts of drought on treatment adherence among people living with HIV in rural South Africa: A qualitative analysis. Climate Risk Management 2022; 36. DOI:10.1016/j.crm.2022.100423.

9 Saunders MJ, Boccia D, Khan PY, et al. Climate change and tuberculosis: an analytical framework. The Lancet Respiratory Medicine 2026; 14: 267–80.

10 StatsSA. Mortality and causes of death in South Africa, 2023: Findings from death notification. Statistics South Africa, 2026.

11 Groenewald P, Nannan N, Joubert JD, editors. South African national cause-of-death validation project: agreement and corrected cause-specific profiles based on data linkage. Cape Town: South African Medical Research Council, 2024.

12 Chersich MF, Wright CY. Climate change adaptation in South Africa: a case study on the role of the health sector. Globalization and Health 2019; 15. DOI:10.1186/s12992-019-0466-x.

13 Barron P, Asia B. The district health system. South African Health Review 2001. DOI:https://journals.co.za/doi/epdf/10.10520/EJC35367.

14 Day C, Gray A, Cois A. Universal Health Coverage – the service coverage index at district level. HST, 2018 https://unstats.

15 Day C, Gray A, Cois A, Ndlovu N, Massyn N, Boerma T. Is South Africa closing the health gaps between districts? Monitoring progress towards universal health service coverage with routine facility data. BMC Health Services Research 2021; 21: 194.

16 Malakoane B, Heunis JC, Chikobvu P, Kigozi NG, Kruger WH. Public health system challenges in the Free State, South Africa: a situation appraisal to inform health system strengthening. BMC Health Services Research 2020; 20: 58.

17 Malakoane B, Heunis JC, Chikobvu P, Kigozi NG, Kruger WH. Improving public health sector service delivery in the Free State, South Africa: development of a provincial intervention model. BMC Health Services Research 2022; 22: 486.

18 Andrade LDMB, Guedes GR, Noronha KVMDS, Santos E Silva CM, Andrade JP, Martins ASFS. Health-related vulnerability to climate extremes in homoclimatic zones of Amazonia and Northeast region of Brazil. PLoS ONE 2021; 16: e0259780.

19 Cutter SL, Boruff BJ, Shirley WL. Social Vulnerability to Environmental Hazards. Social Science Quarterly 2003; 84: 242–61.

20 Muleia R, Maúre G, José A, et al. Assessing the Vulnerability and Adaptation Needs of Mozambique’s Health Sector to Climate: A Comprehensive Study. International Journal of Environmental Research and Public Health 2024; 21. DOI:10.3390/ijerph21050532.

21 Schneiderbauer S, Zebisch M. The Vulnerability Sourcebook: Concept and guidelines for standardised vulnerability assessments. 2014 https://www.researchgate.net/publication/281430219.

22 WHO. Protecting health from climate change: vulnerability and adaptation assessment. World Health Organization, 2014.

23 Davis-Reddy CL, Hilgart A, Hlanane K, Pienaar M, Wilson H, Chiloane L. South African Risk and Vulnerability Atlas. 2020.

24 van Niekerk W, Pieterse A, Le Roux A. Introducing the Green Book: A practical planning tool for adapting South African settlements to climate change. Town and Regional Planning 2020; 77: 103–19.

25 Weaver, Zachariah. Mapping the potential risk of climate change to South Africa’s health sector. 2022.

26 OECD. Handbook on constructing composite indicators : methodology and user guide. OECD, 2008.

27 Greco S, Ishizaka A, Tasiou M, Torrisi G. On the Methodological Framework of Composite Indices: A Review of the Issues of Weighting, Aggregation, and Robustness. Social Indicators Research 2019; 141: 61–94.

28 UNECE. Guidelines on producing leading, composite and sentiment indicators. Inted Nations Economic Commission for Europe : United Nations, 2019.

29 Smits J, Huisman J. The GDL Vulnerability Index (GVI). Soc Indic Res 2024; 174: 721– 41.

30 Ndlovu N, Padarath A. District Health Barometer 2022/23. Durban: Health Systems Trust. Health Systems Trust, 2024.

31 Wang P, O’Brien F, Son J-Y, et al. An updated modeling framework and sensitivity analysis of methodology for the climate health vulnerability index. Nat Commun 2026; 17: 1417.

32 Global Adaptation Initiative Country Index ND-GAIN. University of Notre Dame.

33 Hernangómez D. Using the tidyverse with terra objects: the tidyterrapackage. JOSS 2023; 8: 5751.

34 R Core Team. R: A Language and Environment for Statistical Computing. 2024.

35 Silge J, Robinson D. tidytext: Text Mining and Analysis Using Tidy Data Principles in R. JOSS 2016; 1: 37.

36 Wickham H. ggplot2: Elegant Graphics for Data Analysis, 2nd ed. 2016. Cham: Springer International Publishing : Imprint: Springer, 2016 DOI:10.1007/978-3-319-24277-4.

37 Wickham H, Averick M, Bryan J, et al. Welcome to the Tidyverse. JOSS 2019; 4: 1686.

38 Le Maitre D, Kotzee I. Green Book: The impact of climate change on flooding. Pretoria: CSIR, 2019.

39 Krunoslav K. Social Vulnerability Assessment Tools for Climate Change and Disaster Risk Reduction. UNDP.

40 Schmeltz MT, Marcotullio PJ. Examination of Human Health Impacts Due to Adverse Climate Events Through the Use of Vulnerability Mapping: A Scoping Review. IJERPH 2019; 16: 3091.

41 Estoque RC, Ishtiaque A, Parajuli J, Athukorala D, Rabby YW, Ooba M. Has the IPCC’s revised vulnerability concept been well adopted? Ambio 2023; 52: 376–89.

42 Climate Change and Health Vulnerability and Adaptation Assessment. Geneva: World Health Organization, 2021.

43 LDC Expert Group. The NAP Technical Guidelines. UNFCCC, 2025.

