## Supplementary material for "District-level HIV and TB health system vulnerability to climate and weather hazards in South Africa: a composite index approach"

#### 1 Summary statistics

##### *Table 1: Summary statistics of hazard and vulnerability indicators*

| *Indicator* | *Mean (SE)* | *Range* | *District (min)* | *District (max)* |
|---|---|---|---|---|
| ***Climate and EWE hazards*** | | | | |
| *Heavy rain days* | 3·81 (0·41) | (0, 12·3) | Nelson Mandela Bay | eThekwini |
| *UTCI* | 93 (6·12) | (20·4, 184·4) | Nelson Mandela Bay | Z.F. Mgcawu |
| *Temperature trend* | 0·04 (0·00) | (0·02, 0·05) | Buffalo City | City Of Tshwane |
| *Flood hazard* | 0·07 (0·00) | (0·05, 0·1) | Nelson Mandela Bay | Alfred Nzo |
| *Natural disasters* | 3·04 (0·39) | (0, 10) | Dr Kenneth Kaunda | O.R. Tambo |
| ***Health*** | | | | |
| *HIV mortality* | 44·81 (3·18) | (3, 94) | Ekurhuleni | John Taolo Gaetsewe |
| *TB mortality* | 57·31 (2·92) | (20, 101) | City Of Johannesburg | Chris Hani |
| *HIV prevalence (%)* | 12·25 (0·53) | (3·2, 19·2) | Namakwa | Ugu |
| *Life years lost due to HIV and TB (%)* | 23·53 (0·62) | (13·1, 32·7) | Namakwa | uMkhanyakude |
| ***Health Systems*** | | | | |
| *On ART* (%)* | 68·34 (1·66) | (38·1, 90·7) | West Coast | Xhariep |
| *TB treatment success (%)* | 78·04 (0·9) | (59·8, 89·7) | John Taolo Gaetsewe | West Rand |
| *Hospital beds* | 50·4 (1·78) | (15·3, 75·2) | Bojanala | Mangaung |
| *Medical practitioners* | 33·75 (2·37) | (7·4, 84·7) | Z.F. Mgcawu | Buffalo City |
| *Nurses* | 148·1 (7·24) | (54·4, 296·6) | Z.F. Mgcawu | Buffalo City |
| *PHC* | 1·98 (0.06) | (1, 3·2) | City Of Tshwane | uMkhanyakude |
| *HIV testing (%)* | 37·07 (1·69) | (16·7, 69·3) | Fezile Dabi | Harry Gwala |
| *Health facility density* | 9·98 (0·87) | (2·35, 39·61) | City Of Tshwane | Namakwa |
| ***Human development – adaptation*** | | | | |

| | | | | |
|---|---|---|---|---|
| *Higher education (%)* | 7 (0·38) | (4, 16) | Dr Ruth S. Mompati | City Of Cape Town |
| *Piped water (%)* | 52·75 (2·78) | (22·1, 87·1) | Alfred Nzo | Cape Winelands |
| *Paved roads (%)* | 76 (1·75) | (46, 96) | Z.F. Mgcawu | Ekurhuleni |
| *Access to information (%)* | 96 (0·27) | (89, 98) | Pixley Ka Seme | Amajuba |
| ***Human development- sensitivity*** | | | | |
| *Disability (%)* | 18 (0·63) | (9, 28) | City Of Johannesburg | Dr Ruth S Mompati |
| *Poverty*[§] | 0·05 (0·01) | (0·01, 0·17) | City Of Cape Town | Alfred Nzo |
| *Travel time (min)* | 178·8 (16) | (23·5, 609·9) | City Of Johannesburg | Z.F. Mgcawu |
| *Dependency ratio (%)* | 53 (1·29) | (37, 74) | City Of Johannesburg | Alfred Nzo |
| *Hunger (%)* | 27 (0·78) | (15, 40) | Central Karoo | Nelson Mandela Bay |
| *Unemployment (%)* | 32 (1·08) | (14, 51) | Cape Winelands | Sekhukhune |

[§]This is a composite indicator itself used as found in Fransman & Yu, 2019.

#### 2 Indicator spatial distribution

The following figures display the spatial mapping by district for the HIV/TB sensitivity, health systems adaptation, socio-economic sensitivity and adaptation.

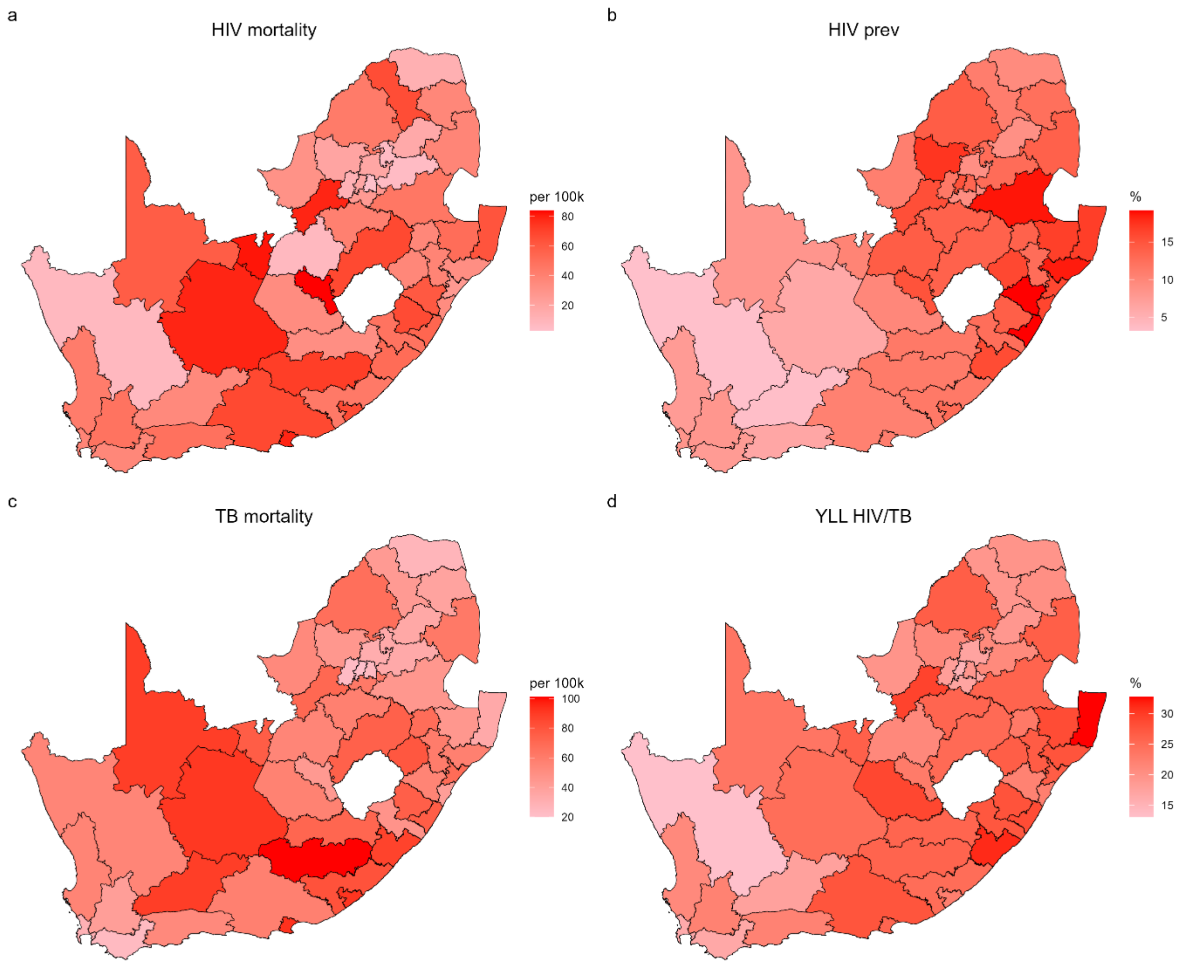

*Figure 1:Spatial distribution of HIV/TB burden*

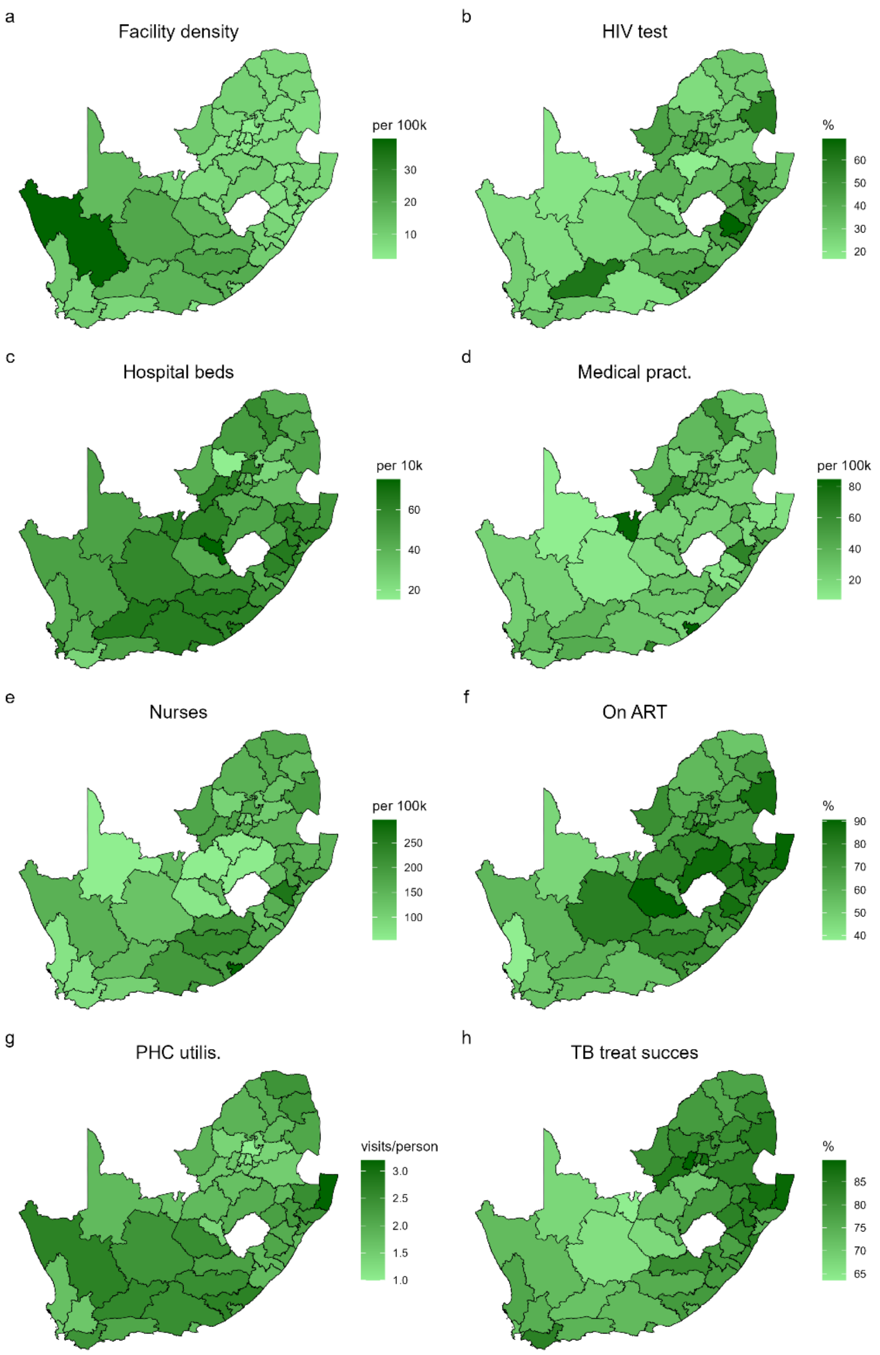

*Figure 2: Spatial distribution of health systems adaptive capacity*

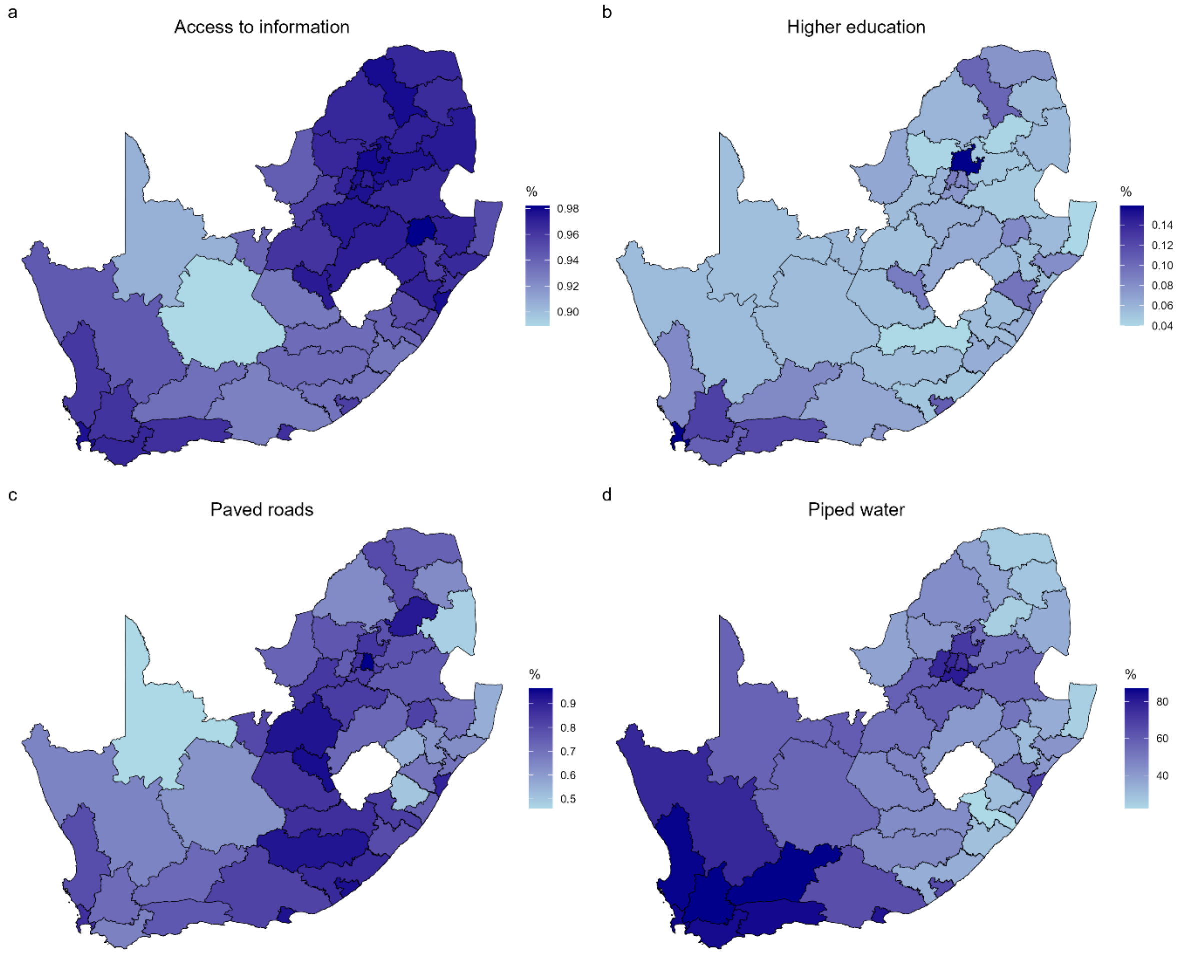

*Figure 3: Spatial distribution of socio-economic adaptation indicators*

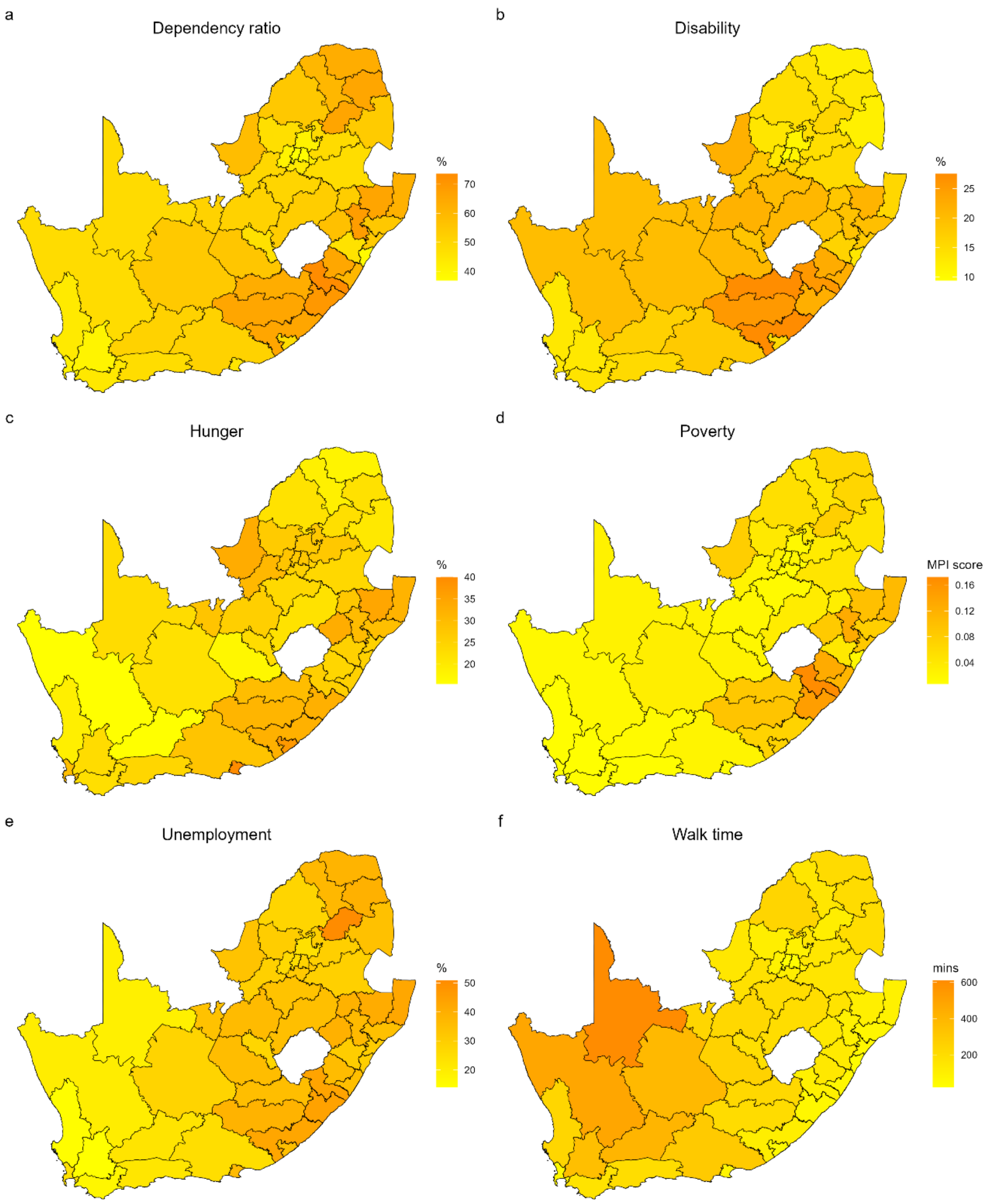

*Figure 4: Spatial distribution of socio-economic sensitivity indicators*

### 3 Indicator correlation structure grouped by dimension

Figure 5 presents the correlation structure among the 27 indicators included in the composite index construction, grouped by dimension. The black outline boxes highlight the within-dimension correlation blocks (diagonal clusters). Our selected indicators exhibit strong clustering within dimensions (e.g., health system access variables correlate with each other; hazard variables correlate with each other). The socio-economic sensitivity indicators (poverty, unemployment, dependency ratio, disability) tend to correlate negatively with socio-economic adaptive capacity indicators (piped water, paved roads, education) which is consistent with susceptibility being inversely related to adaptive resources.

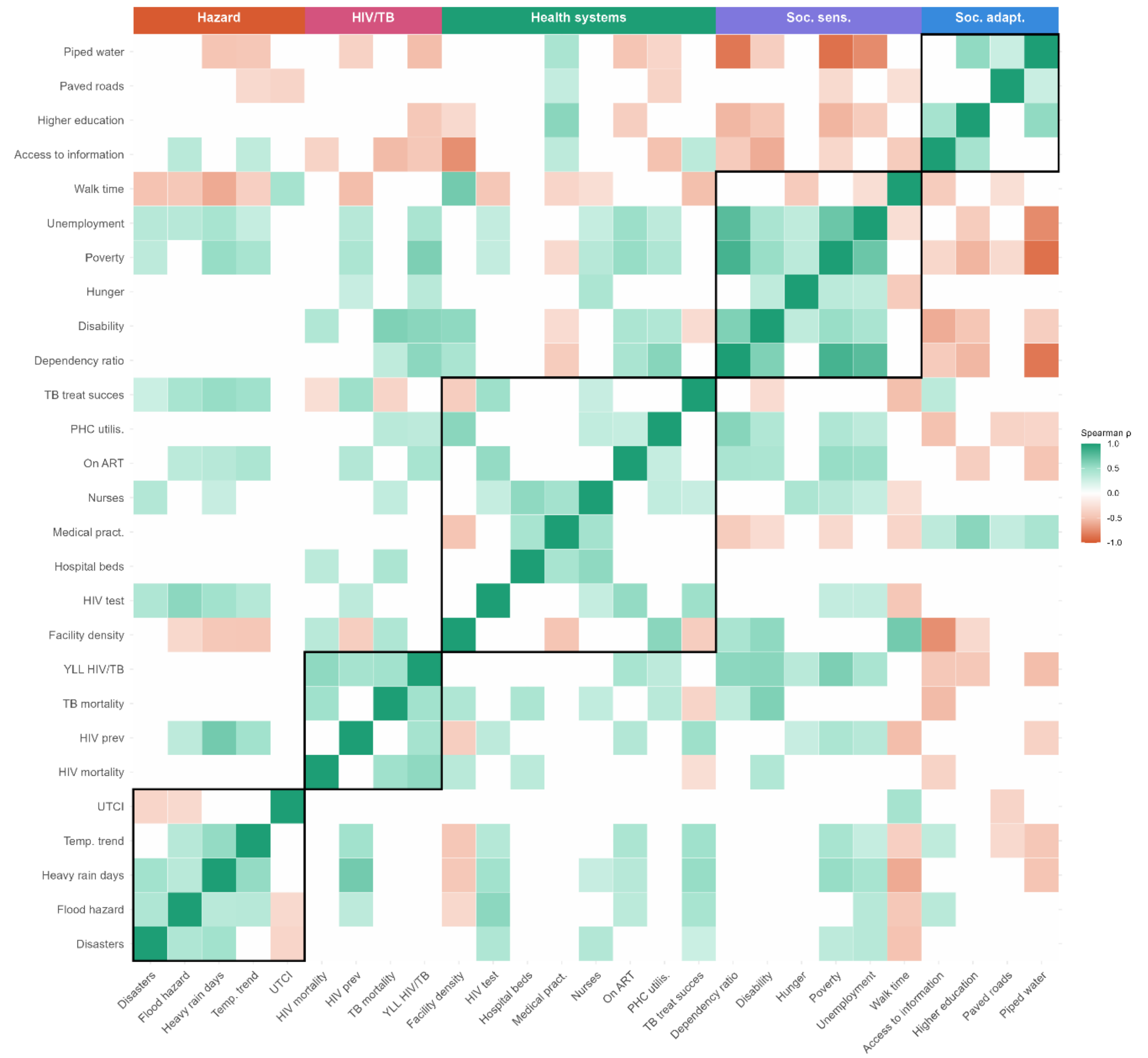

*Figure 5: Correlation structure among the 27 selected indicators. Spearman correlation coefficient ordered by dimension, blank cells denote p-value>0.05*

#### 4 Sensitivity analysis of composite index construction

To assess the robustness of the HIV/TB Vulnerability Index (HVI) and HIV/TB Vulnerability and Hazard Index (HVHI) to methodological choices, we conducted a series of sensitivity analyses examining alternative normalisation and weighting approaches (Tables 2, 3). We also examined how indicator selection influenced district vulnerability rankings by performing a leave-one-out analysis (removing indicators one by one and obtaining the district rankings).

##### 4.1 Normalisation method

The primary analysis used min-max normalisation to transform all indicators to a common scale ranging from 0 to 1 prior to aggregation - a common normalisation method in composite indicator methodology. To evaluate the influence of the normalisation procedure, both HVI and HVHI were recalculated using two alternative methods:

1. Z-score standardisation
2. Percentile rank transformation

For each normalisation approach, dimension scores, component scores, and composite scores were recalculated using the same aggregation structure as the primary analysis. District rankings were then compared with those obtained from the benchmark min-max normalisation. Robustness was assessed using Spearman rank correlation coefficients, Pearson correlations of the resulting index scores, mean and median absolute rank changes, maximum rank changes, and overlap among the ten highest-ranked vulnerable districts (Table 2).

Alternative normalisation procedures had minimal influence on vulnerability rankings. For the HVI, Spearman correlations were 0.989 for percentile-rank normalisation and 0.995 for z-score standardisation. Mean rank changes were small, averaging 1.6 and 0.9 positions, respectively. Maximum rank shifts were limited to seven positions under percentile-rank normalisation and six positions under z-score standardisation. Nine of the ten most vulnerable districts identified under the benchmark approach remained among the top ten under percentile-rank normalisation, while all ten remained unchanged under z-score standardisation.

The HVHI construction also demonstrated robustness across normalisation choices. Spearman correlations exceeded 0.98 under both alternative normalisation methods, mean rank changes were two positions or fewer, and the ten most vulnerable districts were completely preserved under z-score standardisation and largely preserved under percentile-rank normalisation (9 of 10 districts).

##### 4.2 Weighting method

The primary analysis employed equal weighting, whereby indicators are equally weighted to create dimension scores and dimensions are equally weighted to obtain component scores.

To assess the influence of weighting assumptions, principal component analysis (PCA)-derived weights were estimated separately within each index component. Applying PCA within dimension is not advisable due to the small number of indicators in each dimension. Principal components were subjected to varimax rotation and retained until a predetermined proportion of cumulative variance was explained. Sensitivity analyses were conducted using cumulative variance thresholds of 60%, 70%, 80%, and 90%.

For each threshold, indicator weights were derived from the rotated component loadings and used to calculate weighted component scores. HVI and HVHI scores were then recalculated and compared with the benchmark equal-weight specification. As with the normalisation sensitivity analysis, robustness was evaluated using rank correlations, score correlations, rank shifts, and overlap among the ten most vulnerable districts. Higher correlations, smaller rank shifts, and greater overlap indicate greater robustness to methodological assumptions.

Weighting assumptions had a slightly larger influence on rankings than normalisation choices but still produced highly consistent results (Table 3). Across all PCA-derived weighting schemes, HVI Spearman correlations ranged from 0.968 to 0.984, while HVHI correlations ranged from 0.987 to 0.992. Mean rank changes remained modest, varying between 1.6 and 2.6 positions for the HVI and between 1.5 and 1.8 positions for the HVHI.

The greatest deviation from the benchmark occurred under the most parsimonious PCA specification (60% cumulative variance explained), where the maximum HVI rank shift reached 12 positions and 8 of the benchmark top-10 vulnerable districts were retained. However, even under this specification, the HVHI remained highly stable, with a Spearman correlation of 0.987 and complete preservation of the top-10 vulnerable districts. Increasing the PCA variance threshold from 70% to 90% had little additional effect on rankings. Across these specifications, HVI correlations remained above 0.98 and HVHI correlations above 0.99, while 9-10 of the benchmark top-10 districts were consistently identified as highly vulnerable. Mean rank changes remained below two positions for both indices.

*Table 2: Robustness of district rankings for HVI: comparison of alternative normalisation and weighting specifications against the benchmark method.*

| **Alternative specification** | **HVI** | | | | | |
|---|---|---|---|---|---|---|
| | **Spear. r** | **Pears. r** | **Mean rank shift** | **Median rank shift** | **Max rank shift** | **Top-10 overlap** |
| | **Normalisation** | | | | | |
| Percentile rank | 0.989 | 0.990 | 1.6 | 1.0 | 7.0 | 9 |
| Z-score | 0.995 | 0.999 | 0.9 | 0.0 | 6.0 | 10 |
| | **Weighting** | | | | | |
| PCA weights 60% | 0.968 | 0.978 | 2.6 | 1.5 | 12.0 | 8 |
| PCA weights 70% | 0.980 | 0.987 | 1.7 | 1.0 | 12.0 | 9 |
| PCA weights 80% | 0.984 | 0.988 | 1.7 | 1.0 | 9.0 | 9 |
| PCA weights 90% | 0.984 | 0.988 | 1.6 | 1.0 | 10.0 | 9 |

*Table 3:Robustness of district rankings for HVHI: comparison of alternative normalisation and weighting specifications against the benchmark method.*

| | **HVHI** | | | | | |
|---|---|---|---|---|---|---|
| **Alternative specification** | **Spearman r** | **Pearson r** | **Mean rank shift** | **Median rank shift** | **Max rank shift** | **Top-10 overlap** |

| | Normalisation | | | | | |
|---|---|---|---|---|---|---|
| | 0.983 | 0.983 | 2.0 | 2.0 | 8.0 | 9 |
| Percentile rank | 0.998 | 0.999 | 0.7 | 0.0 | 3.0 | 10 |
| Z-score | **Weighting** | | | | | |
| | 0.987 | 0.986 | 1.8 | 1.5 | 6.0 | 10 |
| PCA weights 60% | 0.991 | 0.992 | 1.5 | 1.0 | 6.0 | 10 |
| PCA weights 70% | 0.991 | 0.992 | 1.5 | 1.0 | 6.0 | 10 |
| PCA weights 80% | 0.992 | 0.993 | 1.5 | 1.0 | 6.0 | 9 |
| PCA weights 90% | | | | | | |

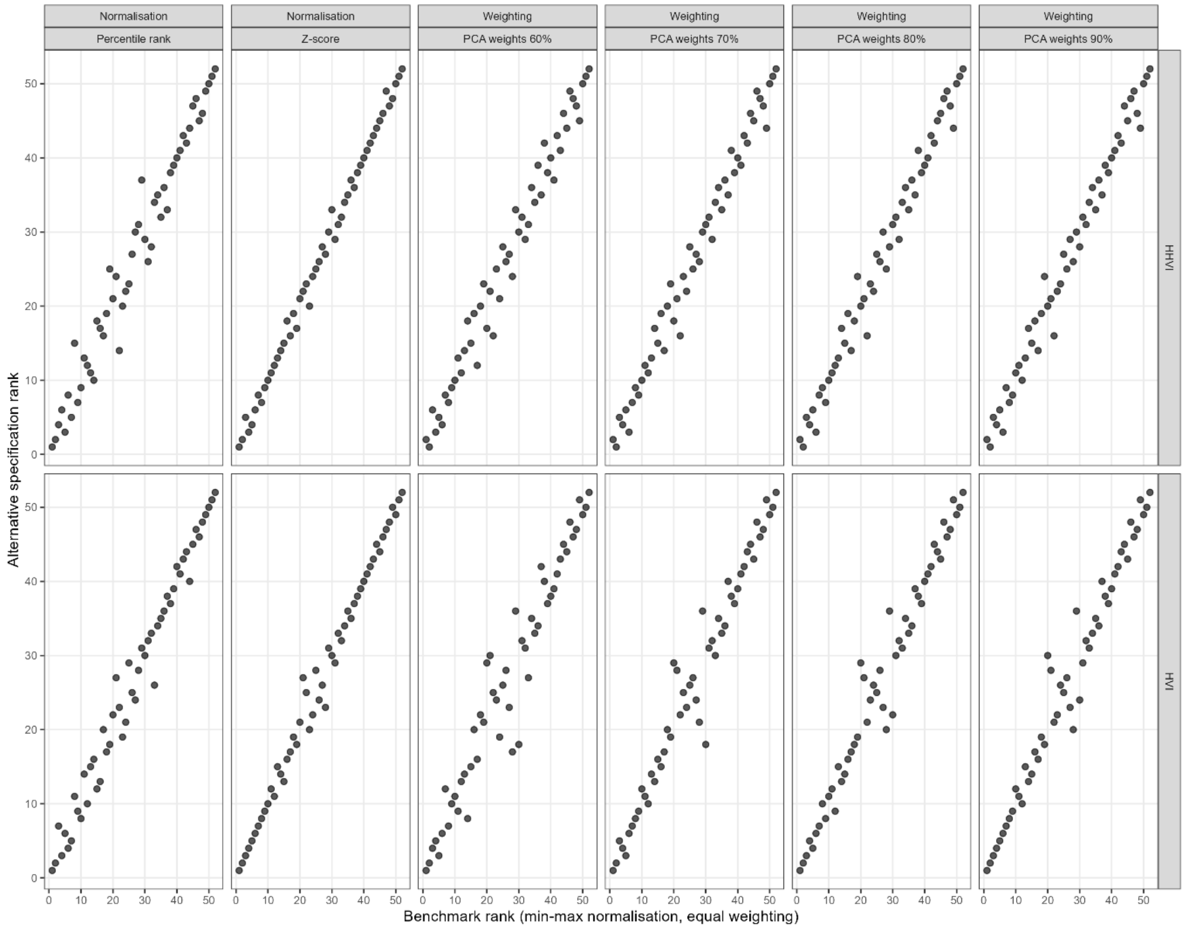

*Figure 6:District ranking stability under alternative methodological specifications. X-axis represents the index scores using the benchmark deductive model and Y-axis represents the index scores using the benchmark inductive model.*

#### 4.3 Leave-one-out analysis

A leave-one-indicator-out analysis is performed to evaluate framework robustness. The method shows how sensitive the final scores and rankings are to the conceptual inclusion of specific dimension indicators. This iterative process systematically evaluates the impact of each individual indicator on the final district rankings. The sensitivity of the framework to each omitted variable is quantified by calculating the mean absolute error, otherwise mean rank change, that occurs when leaving out one indicator. A high mean rank change indicates that the sub-indicator is a primary driver of the index's variance and disproportionally drives the composite indicator. Figure 7 presents the mean rank change by indicator left-out for both vulnerability and vulnerability-hazard indicators. The results signify overall stability and absence of extreme volatility with a mean rank change of 1.2 and a maximum rank change of 2 in the HIV/TB vulnerability index. Moreover, the top ten most vulnerable districts show substantial stability (8-10 districts retained as vulnerable across all permutations).

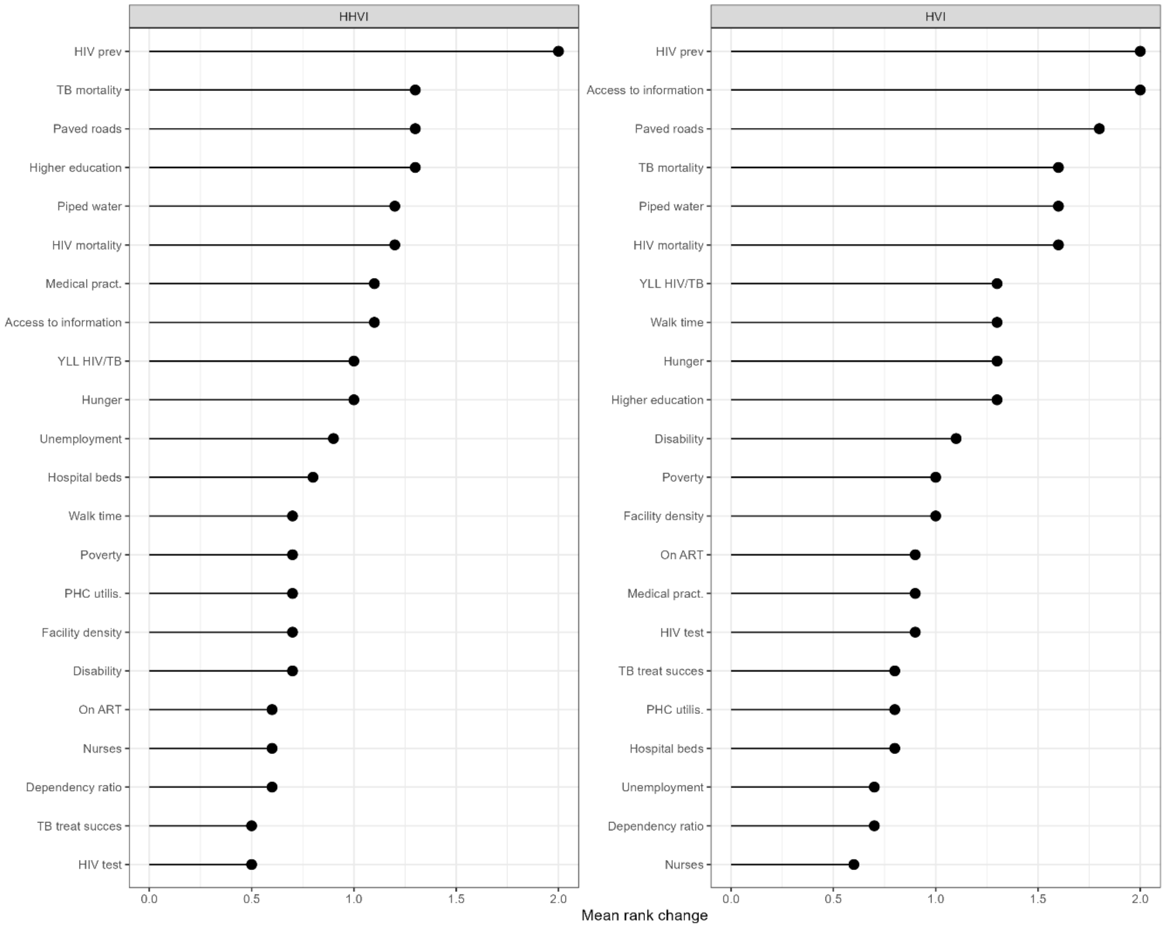

*Figure 7: Mean rank change when the indicators on y-axis is removed from the composite indicator.*

#### 5 Dimension and indicator contributions

This section provides an additional exploration of what drives vulnerability in the top 10 most vulnerable districts. Figure 8 shows the intermediate dimension scores for the ten highest HVHI scoring districts. Most vulnerable districts are characterised by high HIV/TB sensitivity, while they also present low socio-economic adaptive capacity scores, often lower than their health systems adaptive capacity. Climate and weather hazards interact with the districts' overall sensitivity and drive a high HVHI score in already vulnerable districts.

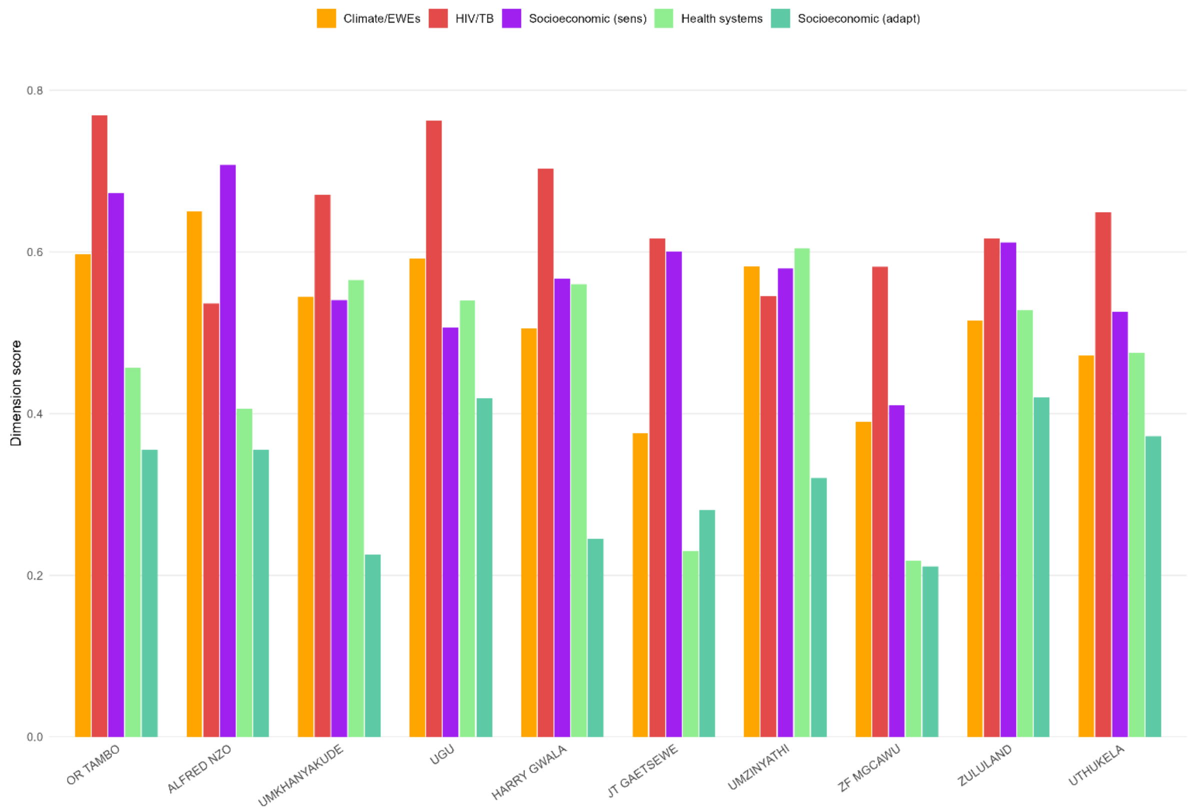

*Figure 8: Dimension score of the 10 highest HVHI scoring districts ordered from highest to lowest score.*

A more detailed picture of what drives vulnerability can be obtained when looking at the indicator heatmap within dimension (Figure 9). Among the ten highest HVHI-scoring districts, a few indicator-level patterns stand out beyond the general dimension-level trends. Z.F. Mgcawu and John Taolo Gaetsewe districts, both in North-West province show a hazard profile driven almost entirely by extreme heat stress (UTCI), rather than disasters, floods, or rainfall. The sensitivity of Z.F. Mgcawu district is dominated by extraordinary long travel time to the nearest health facility showing the challenges in accessing health care in large and sparcely populated districts with poor infrastructure and road systems. Additionally, Z.F. Mgcawu district is characterised by extremely low adaptive capacity in its health systems which fails to offset it sensitivity and lower overall vulnerability.

Umkhanyakude district in KwaZulu-Natal province is affected by both high temperatures and relatively strong rainfall and is deemed vulnerable due to the high socio-economic sensitivity and equally low socio-economic adaptive capacity. It presents notably strong health-system service delivery, especially on ART coverage, PHC utilisation, and TB treatment success rate, although lagging on capacity (medical practitioners and nurses). It's overall high vulnerability is also due to high HIV prevalence and mortality.

Together, these cases suggest that there is no broadly consistent susceptibility pattern that would apply to all districts, and any policy recmmendations and changes have to be made at the district level to address particular shortcomings and strengthen aspects that appear weaker than others.

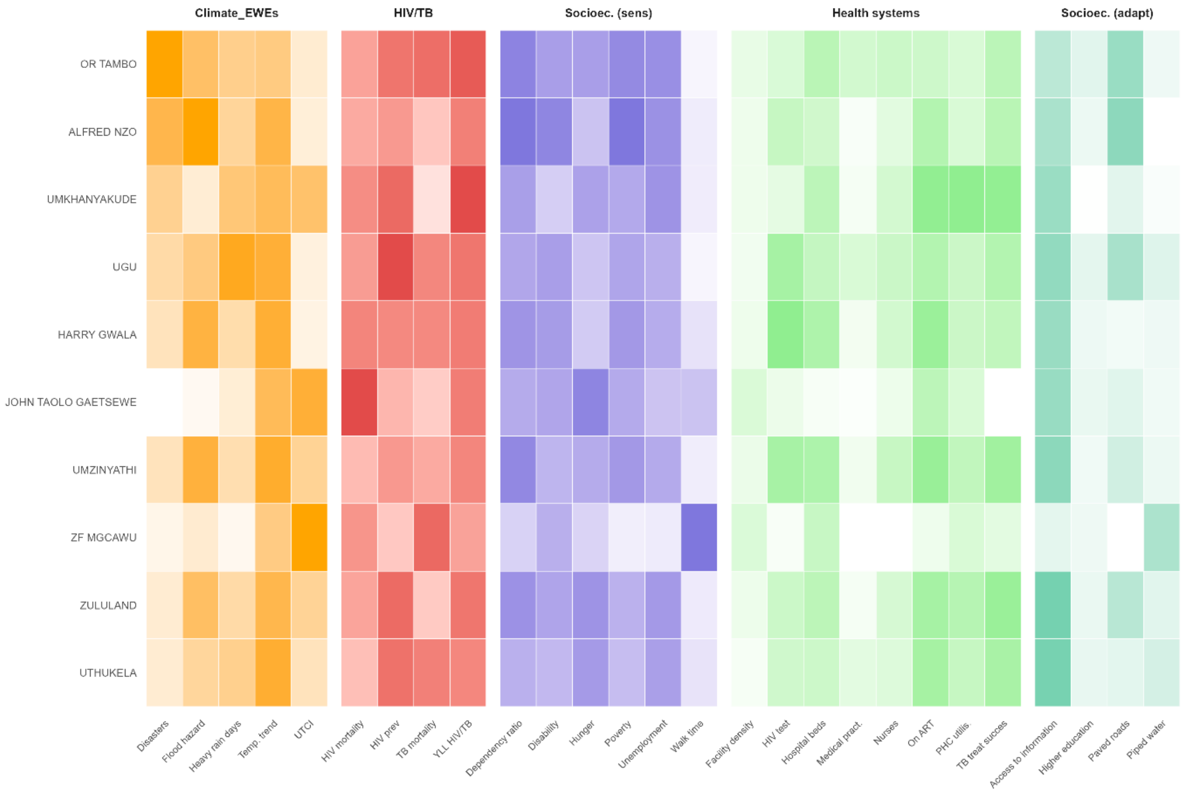

*Figure 9: Heatmap of the 27 selected indicators for the ten highest HVHI scoring districts. Within each dimension, darker shades indicate higher indicator values, with a separate colour gradient used for each dimension.*
